# Menstrual cycle phase and its association with COVID-19 vaccine side effects and subsequent infection: A study of period tracking app users

**DOI:** 10.1101/2025.11.26.25341088

**Authors:** Poppy Alexandra Cooper, Emily R. Boniface, Blair G. Darney, Alison Edelman, Rebecca Sear, Amanda A. Shea, Sarah Walters, Kirsten Weber, Virginia J. Vitzthum, Alexandra Alvergne

**Affiliations:** LSHTM (London School of Hygiene & Tropical Medicine); OHSU (Oregon Health & Science University); Brunel University; Clue (BioWink GmbH, Berlin, Germany); University of British Columbia, Vancouver, Canada; Indiana University, Bloomington, IN, USA; CNRS (Centre National de la Recherche Scientifique), University of Montpellier

**Keywords:** Epidemiology, vaccines, hormones, reactogenicity, follicular phase, menstrual cycle, cycle tracking app

## Abstract

Despite known hormonal influences on immune function, the impact of menstrual cycle phase on vaccine side effects and subsequent infection risk remains unexplored. We used prospectively tracked cycle data from the period tracking app, Clue, matched with an in-app vaccine survey conducted between November 2021 and February 2022. In a sample of 1,474 cycling women, we compared reported side effect and post-vaccination infection outcomes of women vaccinated in the estrogen-dominant follicular phase of their cycle, with those vaccinated in the progesterone-dominant luteal phase. Side effect presence, severity and number were analysed using binary, ordinal, and negative binomial regression models respectively. Post vaccination time to infection (over 67-372 days of follow-up) was compared between the two groups using Wilcoxon rank-sum test and a Cox proportional hazards model.

Follicular-phase vaccination was associated with 35% higher odds of reporting any side effects (OR: 1.35, 95% CI: 1.05–1.66) compared to luteal-phase vaccination. The association remained significant after excluding individuals vaccinated during the bleed phase, and the perimenstrual phase, indicating that hormonal environment, rather than inflammation caused by menstruation or premenstrual symptoms, may drive the association. Median time to infection was 35 days longer for those vaccinated in the follicular phase (Follicular: 200 (140,237) Luteal 165 (107, 2107), p = 0.05), suggesting potential longer-lasting protection from the vaccine.

These findings suggest that menstrual cycle phase should be considered when studying sex-based immune differences and may inform future research on vaccination timing and personalised health strategies.

## Introduction

The coincidence of the COVID-19 pandemic with the rise of big data on menstrual cycles has enabled a substantial leap in studying the previously overlooked relationship between menstrual cycles and vaccine responses ^1,2^. Recent research revealed significant, albeit small and transient, associations between cycle parameters and both COVID-19 vaccination and flu vaccination in numerous populations ^3–9^. Further, prospectively collected data from period-tracker apps indicate that changes in menstrual cycle parameters may be driven by vaccination during the follicular phase ^6,10,11^, reflecting the tight connection between endocrine and immune function ^12,13^. Despite growing interest in menstrual-immune interactions, no research has yet examined whether the timing of vaccination within the menstrual cycle associates with subsequent side effects or infection risk.

Sex differences in vaccine efficacy and side effects have been extensively documented, with these disparities evident even before puberty ^14–20^. Women typically mount stronger humoral and cellular responses to infection and vaccination, and also report more adverse side effects than men^15^. Indications of sex differences in COVID-19 immunity include greater morbidity and mortality in men^21–23^ but more frequent and severe vaccine side effects in women ^24,25^, who also have a higher prevalence of long COVID-19 ^26^. These variations in immune response are linked to behavioural, genetic, and reproductive hormone factors ^27–29^, with reproductive hormones particularly explaining the increased disparity observed during reproductive years ^16,19,30^.

Sex steroid hormones, estradiol (E2) and progesterone (P4), fluctuate over the menstrual cycle, creating additional immune variation beyond baseline sex differences ^13^. While context dependent, estradiol generally enhances immune reactivity in a dose dependent manner ^31^, while progesterone tends to suppress it ^32,33^, creating dynamic variations in immune system functionality across cycle phases. These hormones modulate immune responses through receptors on immune cells including neutrophils, dendritic cells, macrophages and lymphocytes ^16,30,34^. These effects contribute to cycle-dependent variations in susceptibility to infections ^35,36^, manifestations of autoimmune diseases ^37–40^, variation in immune markers of inflammation such as CRP ^41,42^ and total leukocyte count ^43^, as well as specific changes in natural killer and T and B cell responses ^44–46^. Given these hormonal influences on immune function, we can hypothesise how different menstrual cycle phases might affect vaccine responses. The follicular phase—with rising estrogen levels —may enhance both vaccine side effects (reactogenicity) and protection from infection (immunogenicity), while the progesterone-dominant luteal phase may dampen them.

Clarifying the relationship between menstrual cycle phase at vaccination and subsequent vaccine response therefore represents an exciting opportunity to advance personalised medicine, enhancing vaccine effectiveness and individual health outcomes. The transient menstrual disturbances reported following COVID-19 vaccination have raised concerns that could fuel vaccine hesitancy ^2^ making this research particularly timely amid growing recognition of the urgent need for sex-specific and cycle-aware medical research ^13,14,47–51^.

This study investigates whether menstrual cycle phase at the time of first COVID-19 vaccination is associated with vaccine reactogenicity (side effects) and risk of subsequent infection using prospectively collected menstrual cycle data from the period tracking app Clue. Using an integrated dataset of 1,474 individuals who completed a vaccine-focused survey and logged menstrual data prospectively, we tested two primary hypotheses in relation to COVID-19 vaccination: (1) vaccine reactogenicity, measured by the presence, number, and severity of side effects, will be greater when vaccination occurs during the follicular phase; and (2) post-vaccination COVID-19 infection risk will be lower when vaccination occurs during the follicular phase. While cycle-tracking apps have proven to be powerful tools in menstrual health research^9,52–56^, they are an underused resource to examine how vaccine response varies by menstrual cycle phase.

## Results

### Study Population

Following exclusions, 1,474 participants were included for analysis: 760 vaccinated during the follicular phase and 714 during the luteal phase (*Figure 1*). Participant characteristics by vaccination cycle phase are detailed in *Table 1*. The two groups showed no significant differences in demographic characteristics, health behaviours, vaccination details, or survey timing, indicating successful balance between comparison groups.

**Figure 1:**
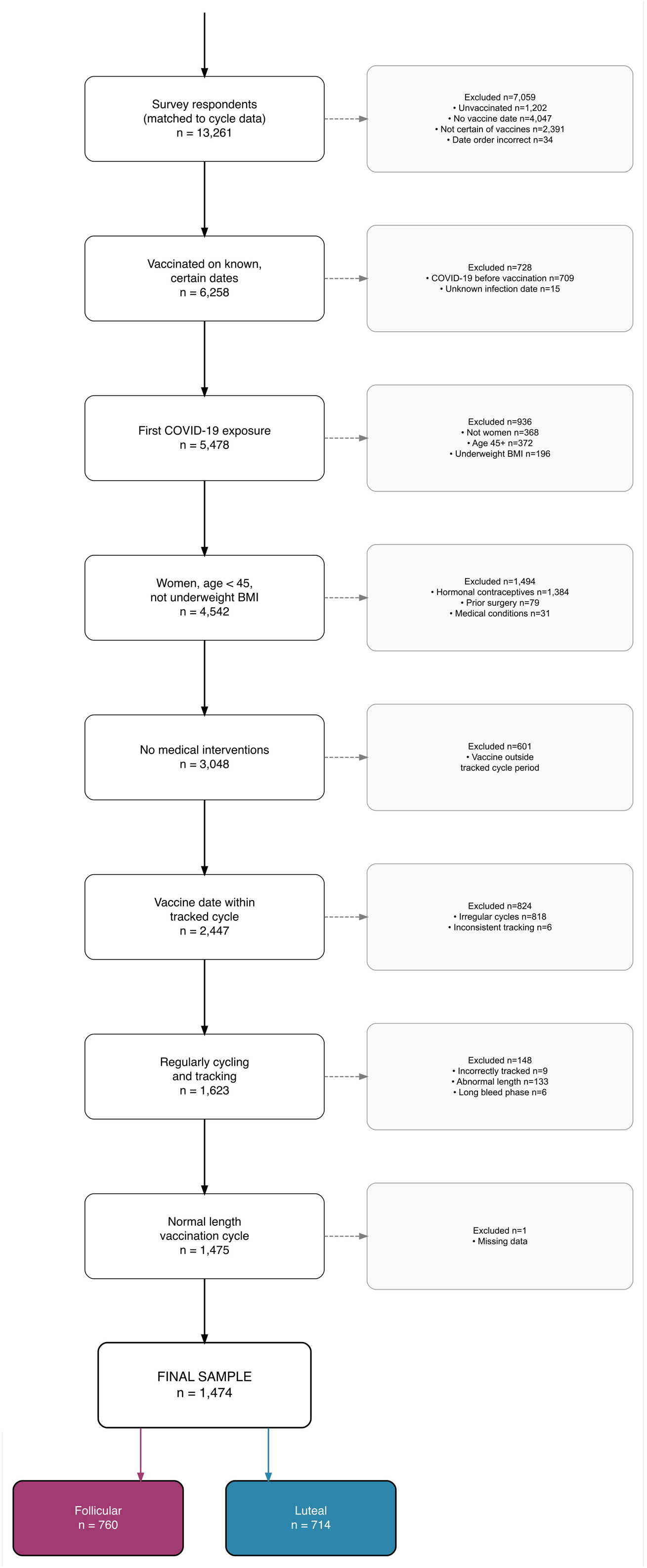
Study Sample Selection

**Table 1.**
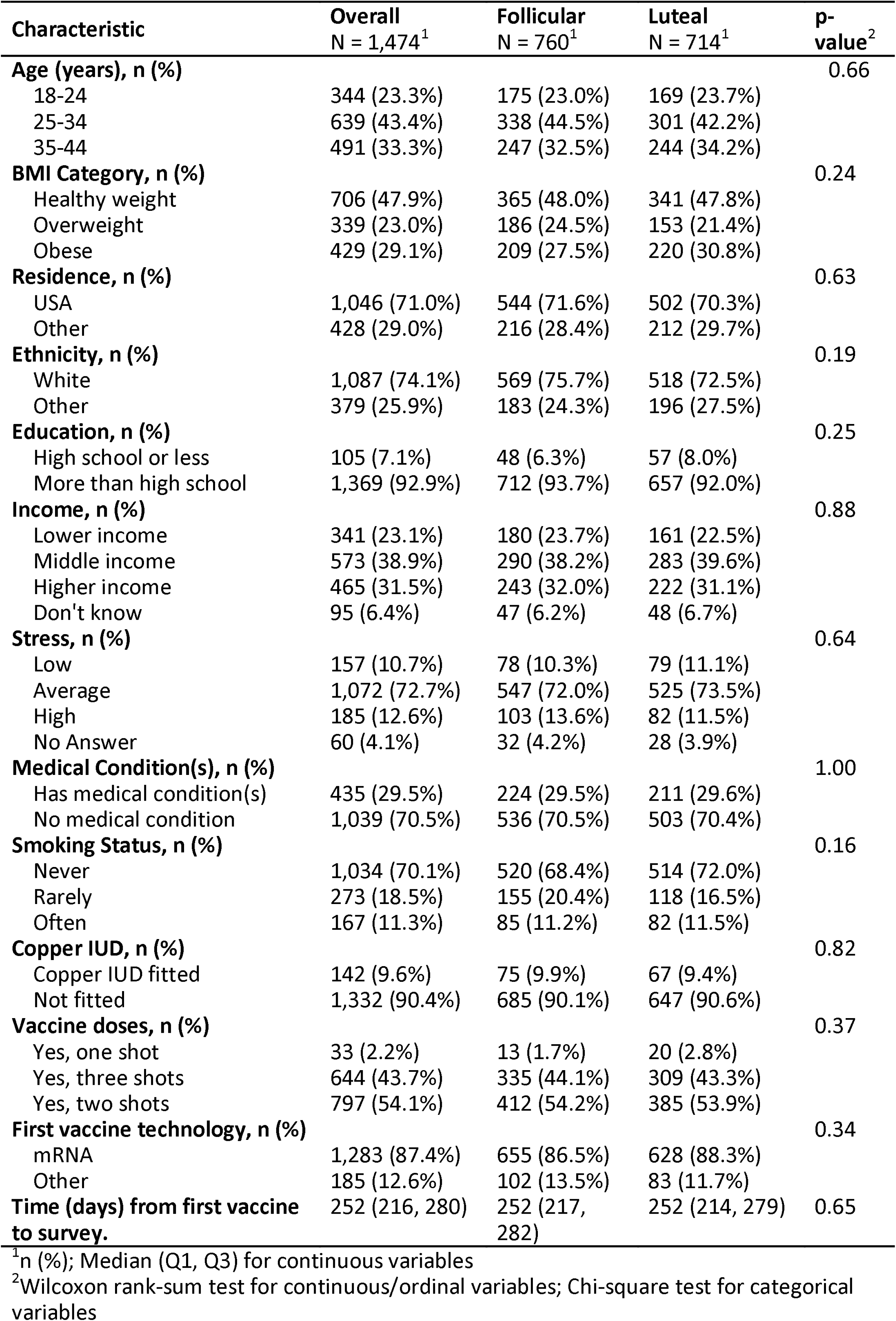
Participant characteristics by menstrual cycle phase (follicular/luteal) at first COVID-19 vaccination.

#### Side Effect Outcomes

Following the first COVID-19 vaccination, a majority (73.2%) of participants experienced side effects, 75.9% following follicular-phase vaccination and 70.3% following luteal-phase vaccination. Most respondents (overall = 48.2%; follicular = 50.8%, luteal = 45.4%, overall p-value = 0.010) reported weak side effects, and the majority (74.9%), reported 3 or fewer side effects (*Table 2*). The most commonly reported side effects were “pain at injection site” (61.5%), “fatigue” (51.0%), and “aches/pains” (35.7%), aligning with previous COVID-19 vaccine studies ^57,58^.

**Table 2.** Side effect outcomes: presence, severity, number and type, by menstrual cycle phase at first COVID-19 vaccination.

| Characteristic | Overall<br>N = 1,474<sup>1</sup> | Follicular<br>N = 760<sup>1</sup> | Luteal<br>N = 714<sup>1</sup> |
| --- | --- | --- | --- |
| **Presence of Side Effects, n (%)** | 1,079 (73.2%) | 577 (75.9%) | 502 (70.3%) |
| **Side Effect Severity, n (%)** |  |  |  |
| None | 395 (26.8%) | 183 (24.1%) | 212 (29.7%) |
| Weak | 710 (48.2%) | 386 (50.8%) | 324 (45.4%) |
| Moderate | 234 (15.9%) | 111 (14.6%) | 123 (17.2%) |
| Severe | 135 (9.2%) | 80 (10.5%) | 55 (7.7%) |
| **Number of Side Effects, n (%)** |  |  |  |
| 0 | 396 (26.9%) | 184 (24.2%) | 212 (29.7%) |
| 1 | 201 (13.6%) | 111 (14.6%) | 90 (12.6%) |
| 2 | 269 (18.2%) | 141 (18.6%) | 128 (17.9%) |
| 3 | 238 (16.1%) | 121 (15.9%) | 117 (16.4%) |
| 4+ | 370 (25.1%) | 203 (26.7%) | 167 (23.4%) |
| **Injection Site Pain, n (%)** | 907 (61.5%) | 478 (62.9%) | 429 (60.1%) |
| **Fatigue, n (%)** | 752 (51.0%) | 412 (54.2%) | 340 (47.6%) |
| **Aches and Pains, n (%)** | 526 (35.7%) | 286 (37.6%) | 240 (33.6%) |
| **Headache, n (%)** | 407 (27.6%) | 218 (28.7%) | 189 (26.5%) |
| **Chills/Fever, n (%)** | 306 (20.8%) | 165 (21.7%) | 141 (19.7%) |
| **Change in Period, n (%)** | 168 (11.4%) | 79 (10.4%) | 89 (12.5%) |
| **Other, n (%)** | 62 (4.2%) | 37 (4.9%) | 25 (3.5%) |
| **Sore Throat, n (%)** | 45 (3.1%) | 22 (2.9%) | 23 (3.2%) |
| **Diarrhoea, n (%)** | 28 (1.9%) | 16 (2.1%) | 12 (1.7%) |
| **Rash, n (%)** | 24 (1.6%) | 8 (1.1%) | 16 (2.2%) |
| **Chest Pain/Pressure, n (%)** | 15 (1.0%) | 10 (1.3%) | 5 (0.7%) |
| **Dry Cough, n (%)** | 14 (0.9%) | 7 (0.9%) | 7 (1.0%) |
| **Difficulty Breathing, n (%)** | 14 (0.9%) | 8 (1.1%) | 6 (0.8%) |
| **Conjunctivitis, n (%)** | 2 (0.1%) | 0 (0.0%) | 2 (0.3%) |
| **Loss of Speech/Movement, n (%)** | 1 (0.1%) | 1 (0.1%) | 0 (0.0%) |
| **Loss of Taste, n (%)** | 1 (0.1%) | 1 (0.1%) | 0 (0.0%) |

<sup>1</sup>n (%); Median (Q1, Q3) for continuous variables

#### Menstrual cycle-phase at vaccination and any reported side effects

Vaccination during the follicular phase was associated with 35% higher odds of reporting side effects compared to luteal-phase vaccination in a model adjusted for age, BMI, smoking status, and pre-existing medical conditions (OR: 1.35, 95% CI: 1.07–1.70). The only other significant predictor of reporting side effects was age, with the youngest participants (aged 18-24) being less likely to report side effects than those aged 25–34 (OR: 0.67, 95% CI: 0.50–0.90; *Figure 2*; *Table 3A*). The association remained robust in sensitivity analyses restricted to participants without factors that might confound the association (no medical conditions, never-smokers, non-obese, White ethnicity, no copper IUD, post-secondary education), with effect sizes ranging from OR = 1.32 to 1.54 (all p≤ 0.05; *Sup. Info. 5*).

**Figure 2:**
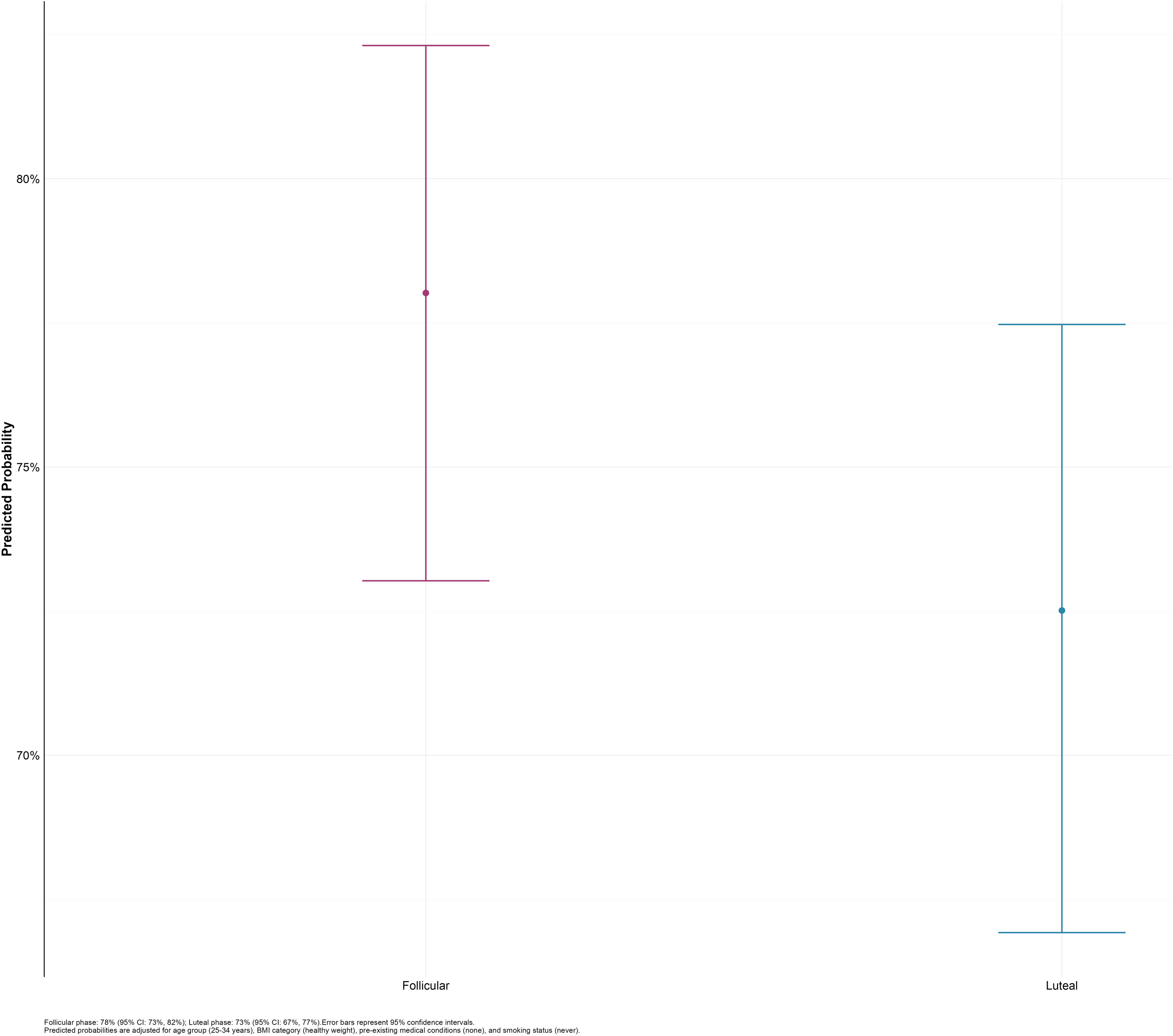
Predicted probabilities of any reported side effects by cycle phase at first vaccination, adjusted for confounders

**Figure 3:**
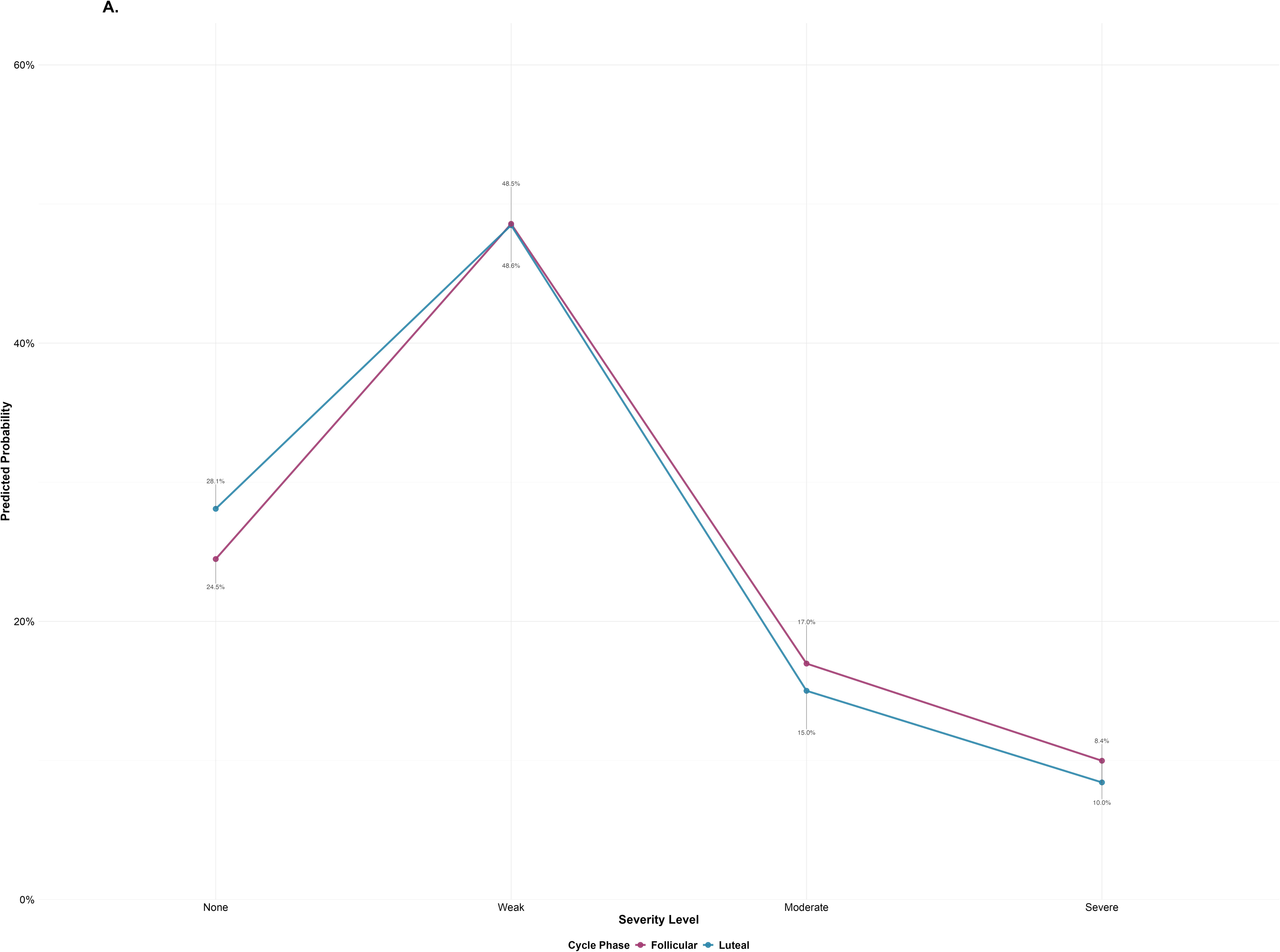
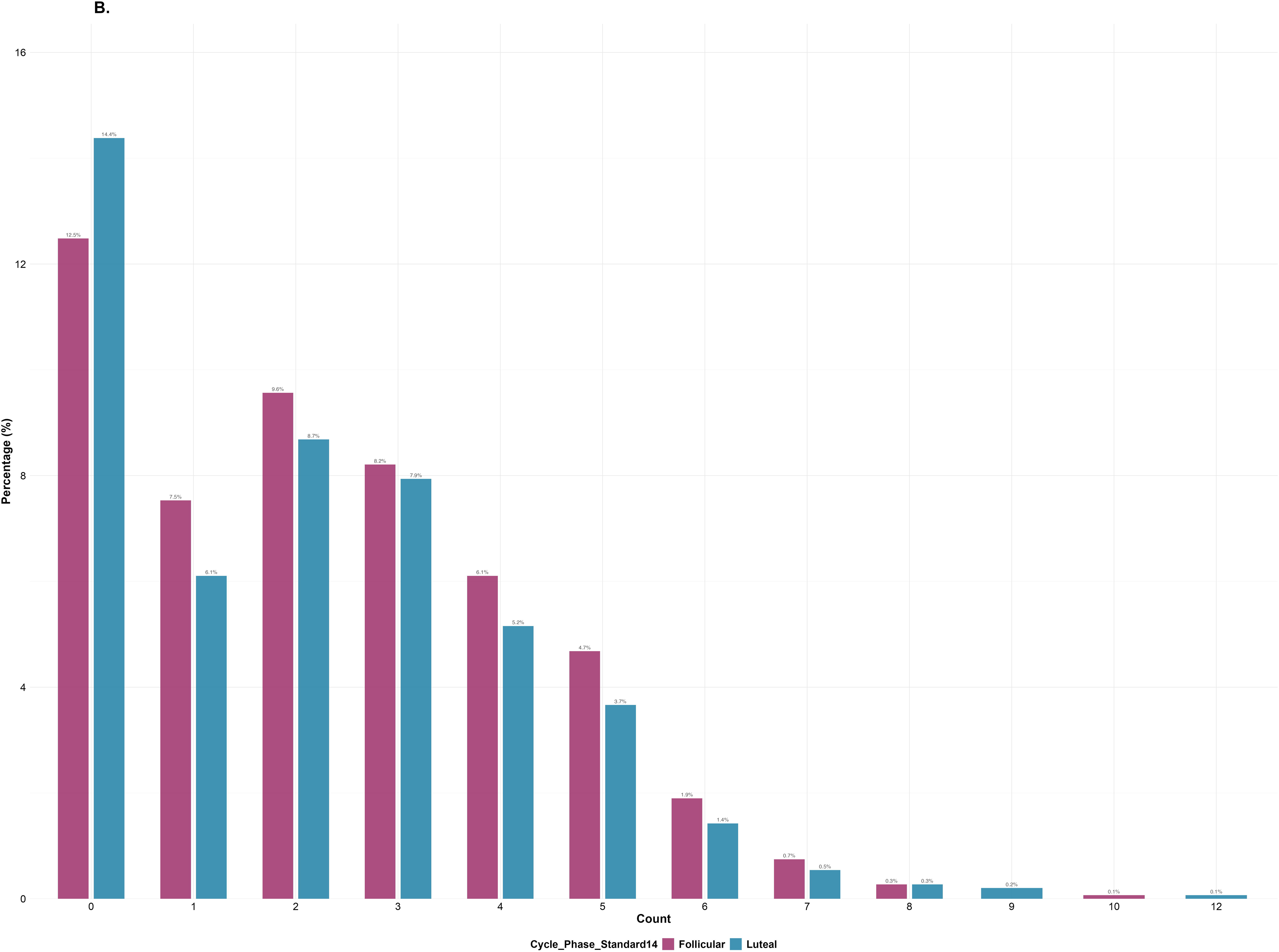
Side effect severity (A) and number (B) by menstrual cycle phase at first COVID-19 vaccination A. Predicted probability of side effect severity by menstrual cycle phase (follicular/luteal) following first COVID-19 vaccination B. Number of side effects by menstrual cycle phase (follicular/luteal at first COVID-19 vaccination

**Table 3:** Side effect presence, severity and number by menstrual cycle phase at first COVID-19 vaccination: Binary logistic, Ordinal logistic and negative binomial regression results.

| Characteristic<sup>1</sup> | A. Side Effect Presence |  |  | B. Side Effect Severity |  |  | C. Side Effect Number |  |  |
| --- | --- | --- | --- | --- | --- | --- | --- | --- | --- |
|  | OR<sup>1</sup> | 95% CI<sup>1</sup> | p-value<sup>1</sup> | OR<sup>2</sup> | 95% CI<sup>2</sup> | p-value<sup>2</sup> | IRR<sup>3</sup> | 95% CI<sup>3</sup> | p-value<sup>3</sup> |
| Cycle Phase (14-day luteal) |  |  |  |  |  |  |  |  |  |
| Luteal | — | — |  | — | — |  | — | — |  |
| Follicular | 1.35 | 1.07, 1.70 | **0.013** | 1.21 | 1.00, 1.46 | 0.056 | 1.08 | 0.98, 1.19 | 0.11 |
| Age Group (per 10 years) |  |  |  |  |  |  |  |  |  |
| 25-34 | — | — |  | — | — |  | 1.11 | 0.98, 1.25 | 0.12 |
| 18-24 | 0.67 | 0.50, 0.90 | **0.007** | 0.76 | 0.59, 0.97 | **0.029** | — | — |  |
| 35-44 | 1.19 | 0.90, 1.58 | 0.2 | 1.15 | 0.92, 1.44 | 0.2 | 1.07 | 0.94, 1.23 | 0.3 |
| BMI Category |  |  |  |  |  |  |  |  |  |
| Healthy weight | — | — |  | — | — |  | — | — |  |
| Obese | 0.94 | 0.71, 1.24 | 0.7 | 0.97 | 0.77, 1.22 | 0.8 | 1.02 | 0.91, 1.14 | 0.8 |
| Overweight | 1.04 | 0.77, 1.41 | 0.8 | 1.09 | 0.85, 1.39 | 0.5 | 1.05 | 0.93, 1.18 | 0.4 |
| Medical Condition(s): Yes (vs No) |  |  |  |  |  |  |  |  |  |
| No medical condition | — | — |  | — | — |  | — | — |  |
| Has medical condition(s) | 0.88 | 0.67, 1.15 | 0.3 | 0.99 | 0.80, 1.23 | >0.9 | 1.00 | 0.90, 1.12 | >0.9 |
| Smoking Status |  |  |  |  |  |  |  |  |  |
| Never | — | — |  | — | — |  | — | — |  |
| Often | 1.28 | 0.87, 1.93 | 0.2 | 1.22 | 0.90, 1.65 | 0.2 | 1.14 | 0.98, 1.33 | 0.083 |
| Rarely | 0.78 | 0.59, 1.05 | 0.10 | 0.81 | 0.63, 1.05 | 0.11 | 0.92 | 0.81, 1.04 | 0.2 |

(A) Binary logistic regression model; <sup>1</sup>OR = Odds Ratio. N = 1474; AIC = 1707.7; BIC = 1755.4; Deviance = 1689.7; Null deviance = 1713.5; Pseudo R<sup>2</sup> = 0.014; Log-likelihood = -844.9
(B) Ordinal regression model; <sup>2</sup>OR = Odds Ratio. N = 1474; AIC = 3587.1; BIC = 3645.4; Residual Deviance = 3565.145; Log-Likelihood = -1782.573; df = 4411
(C) Negative binomial regression model; <sup>3</sup>IRR = Incidence Rate Ratio. N = 1474; AIC = 5779.1; BIC = 5832.1; Deviance = 1775.6; Null deviance = 1787; Pseudo R<sup>2</sup> = 0.006; Log-likelihood = -2879.6; $\theta$ (dispersion) = 2.326; SE( $\theta$ ) = 0.211; Dispersion ratio = 0.881
Abbreviations: CI = Confidence Interval, IRR = Incidence Rate Ratio, OR = Odds Ratio

We conducted sensitivity analyses to eliminate the possibility of vaccine side effects being driven by menstrual or perimenstrual symptoms. After excluding participants vaccinated during menstruation (n=214), the association persisted (OR: 1.34, 95% CI: 1.04–1.72). The association was also confirmed after excluding vaccination in the perimenstrual period (3 days before through 2 days after menstrual onset^59^; n=337; OR: 1.42, 95% CI: 1.10–1.84). We then excluded participants vaccinated during the periovulatory window (days -15 to -12 before menstruation^59^; n=196) to account for distinct hormonal changes around ovulation^60^. The association between follicular-phase vaccination and higher odds of reporting side effects remained (OR: 1.29, 95% CI: 1.00–1.65).

We verified that the cycle-phase association with reporting any side-effects reflected systemic rather than localised side effects by excluding participants who only reported pain at the injection site (the only localised side effect; n=153; *Table 2*).

Follicular-phase vaccination remained associated with increased odds of reporting side effects (OR: 1.33, 95% CI: 1.04–1.68). Finally, we found that younger participants (18-24 years) showed a stronger follicular-phase effect (OR: 1.97, 95% CI: 1.10–3.53) than older age groups (*Sup. Info. 6*). This was driven by lower side effect reporting during the luteal phase in younger participants, while their follicular-phase reporting matched the full sample (*Sup. Info. 14*).

#### Menstrual cycle-phase at vaccination and severity of reported side effects

While individuals vaccinated in the follicular phase showed a trend toward higher odds of reporting more severe side effects compared to those vaccinated in the luteal phase (OR: 1.21, 95% CI: 1.00–1.45), this difference was not statistically significant in the full sample (Table 3B). The only significant predictor of side effect severity was age: individuals in the youngest age group (18–24) had lower odds of experiencing more severe side effects (OR: 0.76, 95% CI: 0.59–0.97) compared to the 25-34 age group.

The positive trend between follicular-phase vaccination and increased severity of reported side effects remained consistent across multiple sensitivity analyses, reaching statistical significance for non-obese respondents (OR: 1.27, 95% CI: 1.01–1.60), never-smokers (OR: 1.29, 95% CI: 1.03–1.62), and non-copper IUD users (OR: 1.24, 95% CI: 1.01–1.51, ; Sup. Info. 7).

We investigated the influence of hormonally distinct parts of the cycle on this association. The effect size and direction remained similar after excluding participants vaccinated during menstruation (OR: 1.21, 95% CI: 0.98–1.49) and the periovulatory window (OR: 1.17, 95% CI: 0.95–1.44). However, when we excluded individuals vaccinated in the perimenstrual period, the effect size increased and the association became statistically significant (OR: 1.29, 95% CI: 1.04–1.61).

To verify whether the observed trend of increased severity following follicular-phase vaccination reflected systemic rather than localised side effects, we excluded participants who only reported pain at the injection site (n=153; Table 2). Follicular-phase vaccination was associated with increased odds of reporting more severe systemic side effects (OR: 1.22, 95% CI: 1.00–1.49).

#### Menstrual cycle-phase at vaccination and number of reported side effects

Follicular-phase vaccination was not associated with the number of reported side effects (IRR: 1.03, 95% CI: 0.94–1.13; *Table 3C*), a finding consistent across sensitivity analyses (*Sup. Info. 9*).

### COVID-19 Infection Outcomes

To test the hypothesis that menstrual cycle phase at vaccination influences subsequent COVID-19 infection risk, we analysed infection outcomes during the observation period (67 to 372 days post-vaccination). A total of 82 of 1,474 participants (5.6%) developed COVID-19 infection post-vaccination. Those vaccinated in the follicular phase had a longer median time to infection (200 days, IQR: 140-237) compared to the luteal phase (164 days, IQR: 107-207; Wilcoxon rank-sum test, p = 0.05, *Table 4, Figures 4A and 4B*).

**Figure 4:**
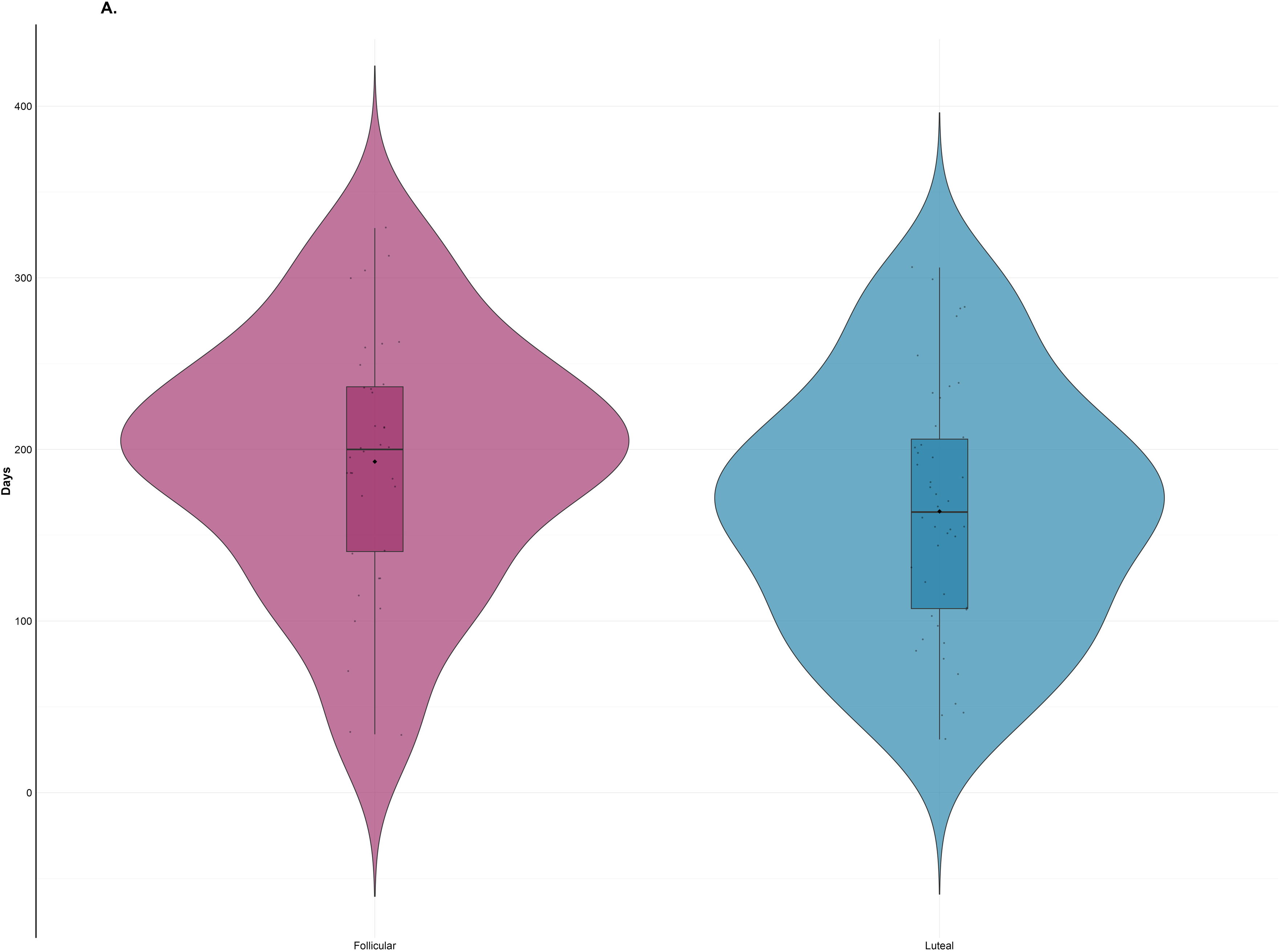
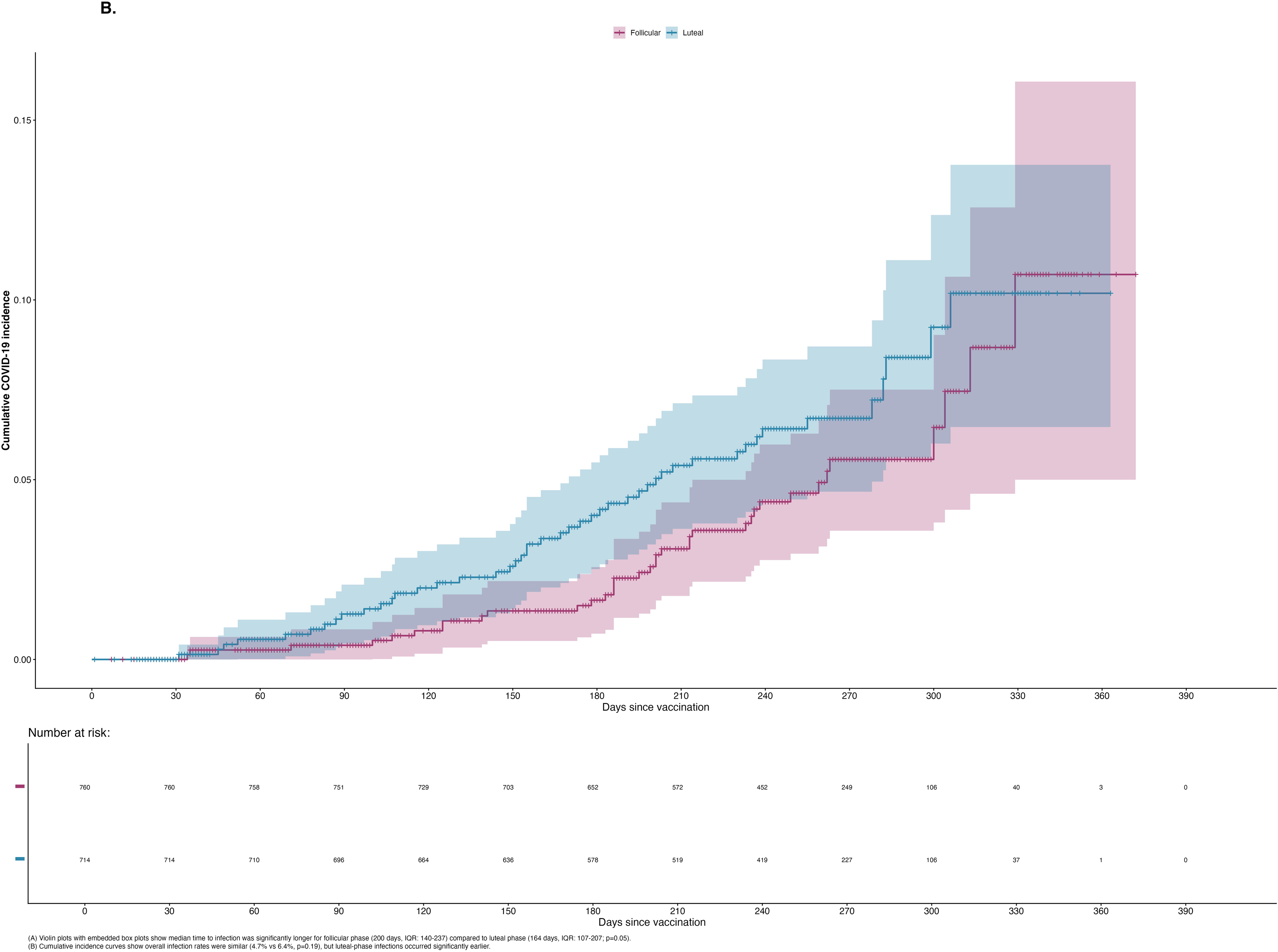
COVID-19 Time to infection (A) and cumulative infection incidence (B) by cycle phase (follicular/luteal) at first COVID-19 vaccination. A. Time to COVID-19 infection by menstrual cycle phase (follicular/ luteal) at first vaccination B. Cumulative COVID-19 incidence by cycle phase (follicular/ luteal) at first COVID-19 vaccination

**Table 4.**
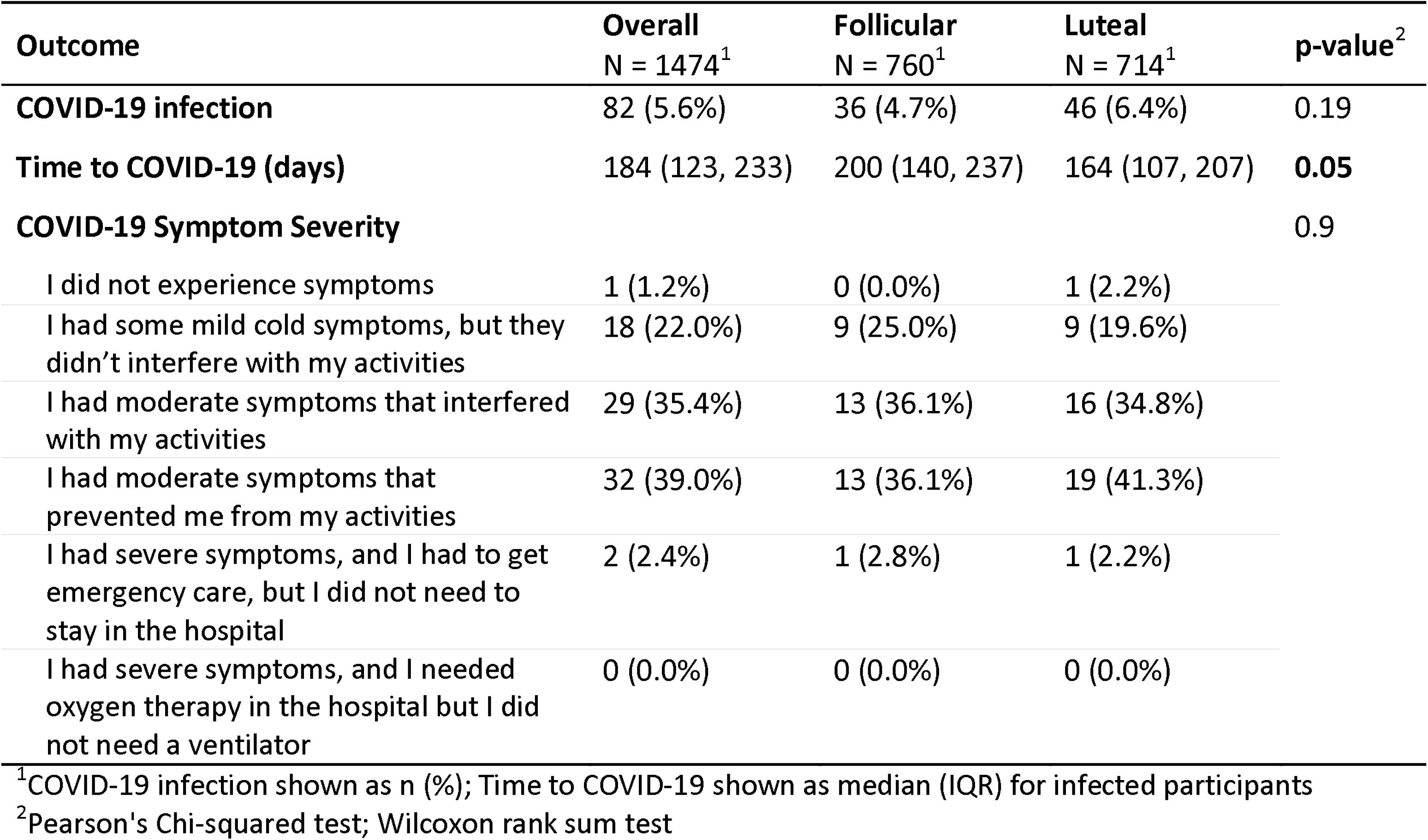
COVID-19 symptom severity post-COVID-19 vaccination by menstrual cycle phase at vaccination.

| Outcome | Overall<br>N = 1474<sup>1</sup> | Follicular<br>N = 760<sup>1</sup> | Luteal<br>N = 714<sup>1</sup> | p-value<sup>2</sup> |
| --- | --- | --- | --- | --- |
| COVID-19 infection | 82 (5.6%) | 36 (4.7%) | 46 (6.4%) | 0.19 |
| Time to COVID-19 (days) | 184 (123, 233) | 200 (140, 237) | 164 (107, 207) | **0.05** |
| COVID-19 Symptom Severity |  |  |  | 0.9 |
| I did not experience symptoms | 1 (1.2%) | 0 (0.0%) | 1 (2.2%) |  |
| I had some mild cold symptoms, but they didn't interfere with my activities | 18 (22.0%) | 9 (25.0%) | 9 (19.6%) |  |
| I had moderate symptoms that interfered with my activities | 29 (35.4%) | 13 (36.1%) | 16 (34.8%) |  |
| I had moderate symptoms that prevented me from my activities | 32 (39.0%) | 13 (36.1%) | 19 (41.3%) |  |
| I had severe symptoms, and I had to get emergency care, but I did not need to stay in the hospital | 2 (2.4%) | 1 (2.8%) | 1 (2.2%) |  |
| I had severe symptoms, and I needed oxygen therapy in the hospital but I did not need a ventilator | 0 (0.0%) | 0 (0.0%) | 0 (0.0%) |  |

<sup>1</sup>COVID-19 infection shown as n (%); Time to COVID-19 shown as median (IQR) for infected participants
<sup>2</sup>Pearson's Chi-squared test; Wilcoxon rank sum test

We used a Cox proportional hazards model to account for censoring, which showed a non-significant trend toward reduced COVID-19 infection risk in participants vaccinated during the follicular versus luteal phase (HR = 0.73, 95% CI: 0.47, 1.13, p = 0.2, *Sup. Info. 11*). The lack of statistical significance is unsurprising given that with only 82 total events; this study had 29% power to detect the observed hazard ratio as statistically significant.

### Adjusting cycle-phase classification methods

Our method for determining cycle-phase at vaccination relied on calendar-based estimation, classifying the last 14 days of the cycle as luteal phase and the remainder as follicular phase, rather than biological confirmation. However, the 14-day luteal phase is not universal: a study of over 600,000 cycles from the Apple Women’s Health Study found the average luteal phase to be 12.4 days, and that it varied by age and cycle length^61^. We therefore tested the associations using alternative luteal-phase lengths (12-day, 13-day, age-adjusted, and cycle-length-adjusted classifications based on that study’s findings). Regardless of classification method, the associations between cycle phase at vaccination and side effect outcomes persisted in the same direction. The 14-day luteal classification consistently provided the best or equivalent model fit across all outcomes and was therefore selected as the primary analysis approach (*Sup. Info. 13*).

## Discussion

Despite 1.8 billion people globally experiencing menstrual cycles each month^62^, historical biases^63,64^, persistent social stigmas and taboos ^65,66^, and practical challenges in data collection^67,68^, mean the menstrual cycle remains understudied. Using prospectively collected menstrual cycle data matched to self-reported survey data on COVID-19 vaccine experiences and infection dates, we found that follicular-phase vaccination was associated with a 35% increased likelihood of reporting side effects and had a non-significant trend towards a longer time to infection (median difference of approximately five weeks) following the first COVID-19 vaccination. We also found that follicular-phase vaccination was associated with a trend toward increased odds of reporting more severe side effects, which reached significance in sensitivity analyses among non-obese respondents, never-smokers, non-copper IUD users, those not vaccinated in their perimenstrual phase, and those who reported systemic side effects. These findings represent preliminary evidence that menstrual cycle phase at the time of vaccination may influence vaccine reactogenicity and warrants further investigation regarding protective efficacy.

Higher rates of any reported side effects when vaccinated during the follicular phase align with this phase’s pro-inflammatory profile ^42^, in contrast to the less pro-inflammatory luteal phase ^13,69^. This interpretation aligns with evidence that symptoms of epilepsy, asthma, rheumatoid arthritis, irritable bowel syndrome, and diabetes fluctuate across the menstrual cycle ^70^. Rising estradiol levels approaching ovulation could promote inflammatory cytokine production ^15,30,33,71^, potentially leading to greater side effect reporting. In contrast, progesterone, which dominates in the luteal phase, can reduce cytokine production ^72–74^. Because inflammation is a crucial part of the innate immune response, these findings may reflect stronger innate immune activation following follicular-phase vaccination. The observation that follicular-phase vaccination increased odds of reporting systemic side effects, a plausible proxy for innate immune response□^9^, and that these were more severe, supports this hypothesis.

Importantly, this association persisted after excluding participants vaccinated during symptom-heavy points of the menstrual cycle: menstruation (the start of the follicular phase), an inflammatory event associated with symptoms such as fatigue, headache, breast tenderness and cramping^75^; peri-menstruation (defined as the last three days of a cycle and first two days of the subsequent cycle^59^), a hormonally distinct window marked by low estrogen and progesterone, also associated with elevated symptom reporting ^59,76^; and ovulation, which follows a peak in estrogen levels, and has been linked to a higher prevalence of symptoms such as of migraines ^77^ and ovulation pain^78^. These sensitivity analyses suggest that our findings are unlikely to result from women misattributing menstrual cycle side effects to the vaccine. Moreover, removing participants vaccinated during peri-menstruation strengthened the association between follicular-phase vaccination and more severe side effects, indicating that any misattribution would have attenuated our finding rather than artificially inflating it.

The association between vaccination in the follicular phase and higher likelihood of reported side effects persisted in white, never-smokers, non-obese, non-copper IUD users and those without any pre-existing medical conditions. More severe side effects were also more likely following follicular-phase vaccination when obese participants, smokers, and copper IUD users were excluded. We had previously excluded hormonal contraceptive users, participants aged 45+, and those reporting irregular cycles (*Figure 1*), so the observed cycle-related differences may reflect cyclical hormone level changes rather than underlying medical conditions^79^, hormonal contraceptive symptoms^80^, low-grade inflammation linked to obesity^81,82^, perimenopause symptoms^83^, copper IUD symptoms^84,85^, or smoking ^86,87, 88^.

Age modified the association between follicular-phase vaccination and reporting any side effects, with the younger age group showing the strongest association. This finding could have several explanations. Most simply, younger women often experience more variable menstrual cycles ^53,61,89^, which could lead to cycle-phase classification errors. If they happen to be misclassified in one direction (e.g. owing to a shorter luteal phase^61^) this might amplify apparent associations. If the observed modification reflects a genuine age-related difference rather than measurement error, the underlying data offer some clues. Younger women in our sample reported side effects less frequently in the luteal phase than the full sample, while their reporting rates in the follicular phase were comparable to the full sample (*Sup. Info. 14*). This could indicate more pronounced progesterone-mediated immunosuppression in the luteal phase among younger women (who are at peak fertility^90^), making the contrast with follicular-phase responses more apparent.

Progesterone’s role in promoting an immunotolerant environment for conception and pregnancy^12^ may be particularly strong in this group, leading to fewer reported side effects. Regardless of the underlying mechanism, the finding underscores the importance of considering age-related differences in hormonal sensitivity when evaluating menstrual cycle phase and vaccine interactions.

Our observation of a 5-week longer time to infection following follicular-phase vaccination in our bivariate analysis may indicate more durable vaccine protection, although reduced infection risk following follicular-phase vaccination was not confirmed in the underpowered, adjusted Cox model. Time to infection and infection risk are appropriate outcome measures, reflecting the non-sterilising protection profile of COVID-19 vaccines compared to those that achieve near-complete sterilising immunity (preventing the virus from establishing infection at all)^91^. COVID-19 vaccines confer lasting protection against severe disease but less complete and often rapidly waning protection against mild or asymptomatic infection^92^. Physiologically, estrogen levels have been associated with enhanced B cell activity and T follicular helper cell responses, both important for long-term immunity, while progesterone, can suppress these responses^30^. The increased odds of side effects, coupled with the potential for longer vaccine protection following follicular-phase vaccination could reflect a careful immune balance. Specifically, the estrogen-dominant follicular phase may support coordinated adaptive immune activation with appropriate inflammatory signalling, while the progesterone-dominant luteal phase may favour tolerance over inflammation and therefore lower reactogenicity and potentially less robust protection.

However, greater vaccine reactogenicity does not necessarily correlate with better disease protection. Previous research on COVID-19 vaccine reactogenicity and protection have been inconsistent^93,94,88^. While our findings suggest a trend toward longer protection following follicular-phase vaccination, the limited number of infections in this study means it was underpowered to determine whether a true difference in protection exists

Understanding whether follicular-phase vaccination is associated with both increased reactogenicity and enhanced protection, or increased reactogenicity alone, has important implications. Reproductive-aged women show greater vaccine hesitancy, with fear of side effects and potential long-term health implications often cited as reasons^95,96^. Clarifying any potential relationship between cycle phase at vaccination, side effects, and subsequent protection could help this population make more informed decisions. Additionally, further research into the already observed associations between vaccination timing and menstrual bleeding patterns and cycle length ^6,10,11^ , as well as potential influence on ovulation timing, could address concerns about menstrual changes and fertility that have been reported as barriers to uptake^2,97^. While COVID-19 vaccines have demonstrated clear effectiveness in reducing infection risk, understanding whether timing vaccination with menstrual cycle phase could optimise individual outcomes is an important avenue for future research.

### Strengths and Limitations

This study draws on a large, international dataset of prospectively collected menstrual cycle data linked to COVID-19 vaccination and infection outcomes. This approach reduces recall bias compared to retrospective self-reporting for cycles. Stringent eligibility criteria and DAG (directed acyclic graph) informed covariate selection, and multiple sensitivity analyses strengthened internal validity. These analyses focused on responses to participants’ first COVID-19 vaccine dose, avoiding potential confounding from cumulative or mixed vaccine exposures. The observed associations are biologically plausible given currently known hormonal effects on immunity, appearing to operate not just between sexes but also within women, depending on cycle timing ^15^.

Several limitations should be acknowledged. Extensive exclusions limit generalisability, especially to those aged 45+, those with irregular cycles, varied gender identities, underweight BMI, prior COVID-19 infection or using hormonal contraceptives. As a self-selected sample of app users, participants differ from the general population in health behaviours, digital literacy, and socioeconomic status. The sample was predominantly white, residing in the USA, highly educated and affluent, which limits generalisability (*Table 1*).

Cycle phase was estimated from tracking data without biological markers; ovulation was inferred rather than measured, and hormone levels were not available. Although excluding irregular cycles likely reduced inclusion of anovulatory cycles associated with poor health, regular cycles can also be anovulatory, and immune responses may differ accordingly^69^. Misclassification is possible due to the assumption of a fixed 14-day luteal phase, despite evidence that length varies between and within individuals^61^. This assumption creates a structural imbalance: participants with longer cycles are classified as follicular phase for a greater proportion of their cycle. Consequently, more vaccination dates fall within the follicular-phase classification than the luteal-phase, potentially introducing confounding if cycle length itself relates to immune responses or if individuals with systematically longer cycles differ in unmeasured ways. We cannot entirely rule out that observed associations reflect, at least partially, underlying differences between individuals with varying cycle lengths rather than solely the direct immunological effects of hormonal fluctuations. Even when phases are correctly classified, hormone patterns can vary considerably between individuals and cycles^97^, which tracking data cannot capture.

Nonetheless, our results were robust across sensitivity analyses that removed menstruation, peri-menstruation and periovulation, symptom heavy sections of the cycle, as well as analyses using different cycle-phase classification methods. The reported model was the best fit to the data (*Sup. Info. 13*). Future studies using biological confirmation of ovulation and cycle phase, combined with direct hormone measurements, would help disentangle these potential sources of confounding.

COVID-19 infection was self-reported and limited to events between vaccination and survey completion (a period ranging from 65–372 days), introducing variability in exposure opportunity and potential recall bias. We also did not have information that would capture differential exposure risk (e.g. working from home vs in-person) during lockdown periods of the pandemic, which could significantly bias the infection results. With n=82 infection events, our model achieved just 29% power to detect a hazard ratio of 0.73 (the effect size observed in our study) at α = 0.05. Our choice of time to infection as an endpoint was determined by available data rather than optimal study design, though it does provide a useful metric for COVID-19 vaccines protection profiles^92^. While direct measurement of antibody kinetics or cellular immune responses across menstrual cycle phases would provide more mechanistic insights, such biomarker data were not available. These preliminary findings warrant replication in larger cohorts with prospective biological sampling to measure immune responses directly, which would clarify both the magnitude of any cycle-phase effects and their clinical significance.

## Conclusion

Effective vaccination requires a balance between immunogenicity, the ability to trigger strong adaptive immunity, and reactogenicity, the risk of excessive inflammation. As sex hormones influence both innate and adaptive immunity, and fluctuate across the menstrual cycle, understanding their role is essential. Our findings offer a compelling case for further research into the relationship between cycle phase at vaccination and vaccine outcomes, using direct measurement of hormonal and immune markers. Beyond menstrual cycles, these findings provide further evidence of the importance of investigating hormonal diversity between and within women, not just between sexes, in immunological research.

## Methods

### Data sources and population

This research received approval from the French National GDPR Committee (CNIL) and the LSHTM Ethics Committee (ref: 31539). The “Period and the Pandemic” survey, conducted among users of the Clue app (BioWink GmbH, Berlin, Germany), collected self-reported data that were linked to participants’ prospectively logged menstrual cycle information. All data were obtained with participants’ consent, and identifying information was removed prior to data transfer and analysis.

Users aged 16 to 58 years old with a registered Clue account, who had consented to the use of their pseudonymised data for research purposes, were sent an in-app message inviting them to take part in the survey. Participants were eligible for inclusion in the analysis if they had experienced menstrual periods or withdrawal bleeds and had not been pregnant or breastfeeding since January 2020.

The survey was conducted between 29 November 2021 and 8 February 2022, and sent to app users in the United States, United Kingdom, Canada, and Australia. It collected demographic data (age, ethnicity, residence), health markers (height, weight, pre-existing conditions, stress), lifestyle factors (smoking, contraceptive use), Covid-19 infection history (test type, test and diagnosis dates, symptoms, symptom onset date), vaccination details (type, date, number of doses—up to three), vaccine side effects (type: 16 options, and severity), and long COVID status (ongoing symptoms). Survey responses were paired, whenever possible, to longitudinal cycle data, including cycle and period start and end dates tracked from three cycles prior to cycle containing respondent’s first reported vaccine date and end of survey.

### Variables and Data Processing

Age was transformed into a categorical variable (18-24, 25-34 and 35-44) to maintain adequate numbers in each group). BMI was calculated using the standard formula and categorised as ‘underweight’, ‘healthy weight’, ‘overweight’ and ‘obese’ according to standard parameters^98^. Ethnicity was reported differently across the four countries, so responses were combined and split into seven categories (“White”, “Black”, “Hispanic”, “Asian”, “Middle Eastern”, “Multiple” and “Other”) and then collapsed into “White” and “Other”, owing to low counts in non-white groups (*Sup. Info. 4*). “Prefer not to answer”, was listed as “Missing” in the Ethnicity variable.

Residence was collapsed to “USA” and “Other”, owing to low counts in other countries. Education was grouped into “high school or below” and “more than high school”. Pre-existing medical conditions (for full list see *Sup. Info. 4*) were recoded to: “Has medical condition(s)”, “No medical conditions” and “prefer not to answer”. A composite smoking frequency score was calculated by numerically recoding tobacco, e-cigarette (vape), and cannabis use responses from 0 (“never or not in past 2 years”) to 4 (“several times a day”), then summing the recoded values across the three substances. The combined score was then capped at 5 and recoded into a categorical variable with three levels: “Never” (0), “Rarely” (1-3), and “Often” (4+).

Income was reported in six local currency categories for each country. These country-specific categories were numerically coded (1–6) to standardise them across UK, USA, Canada, and Australia, then recategorised as “Lower income*”* (1–2), “Middle income (3–4), and “Higher income” (5–6) groups; “Don’t know” responses were included as a separate category and removed for a sensitivity analysis. The administered vaccine was recoded by its technology as “mRNA” (Pfizer-BioNTech/Moderna) or “Other” (Oxford-AstraZeneca/Johnson & Johnson/ Novovax). mRNA technologies accounted for >85% of responses (*Table 1*). Hormonal contraceptive users were excluded and copper IUD users retained, and a binary variable created (“Copper IUD fitted”/ “Not fitted”). Participants reported stress they felt during the pandemic on a 0–10 scale. This variable was recoded with three categories: “Low” (below median minus IQR), “High” (above median plus IQR), or “Average” (all others) based on sample distribution and non-responses listed as “No answer”.

#### Response variables

Vaccine side effects were summarised using three variables: presence (binary), number (count, up to 16), and severity (ordinal). Severity was based on participant-reported interference with daily activities: “no side effects”, “mild side effects that did not interfere with activities”, “symptoms that interfered with activities”, “symptoms that prevented me from my activities” and “symptoms that required emergency care or hospital visit”. These were recoded to “none”, “weak”, “moderate” and “severe”, with “severe” combining the last two levels owing to low counts.

We used time to COVID-19 infection as our primary endpoint for assessing vaccine protection. This measure is appropriate for COVID-19 vaccines given their incomplete protection profile (67-92% efficacy depending on platform^99^) and regular breakthrough infections, unlike vaccines with near-complete protection where binary success/failure outcomes are more appropriate. Time-to-infection allows detection of potential differences in the timing and durability of vaccine-induced protection development across menstrual cycle phases.

Time to infection was calculated from the date of the first vaccine as recorded by survey respondents to the earliest date given for any of COVID-19 symptom onset, COVID-19 positive test date, or COVID-19 positive diagnosis date. COVID-19 infection dates were self-reported by participants during the survey period (67-372 days post-vaccination). COVID-19 infection dates were only provided by month and year, so the 15th day of each reported month was assigned as the infection date to represent the midpoint of the likely infection window.

#### Inferring menstrual cycle phase

We matched reported first vaccine date to corresponding cycle date range and then assigned cycle phase (follicular/luteal) at vaccination. Luteal phase was defined as the 14 days preceding the onset of the next menstrual period, counting backward from the last day of the cycle (the day before bleeding begins) through day −13. All remaining cycle days were assigned to the follicular phase ^70^. Additional classifications were used in sensitivity analyses (see below).

#### Exclusions

Exclusions were applied as detailed in *Figure 1.* A total of 13,261 registered Clue users who responded to the survey were matched to their prospective cycle data. We excluded unvaccinated individuals, those unable to recall or specify their vaccine date, and participants who reported a COVID-19 infection or gynaecological surgery prior to vaccination. Additional exclusions were based on demographics (did not identify as women, age 45+), underweight BMI (owing to low counts), hormonal contraceptive use (Copper IUD users were retained) and a response of “prefer not to answer” regarding pre-existing medical conditions. Participants who had not tracked the cycle in which their first COVID-19 vaccination occurred were also excluded. We defined regular cycles according to FIGO (International Federation of Gynaecology and Obstetrics) criteria (24-38 days, menstruation length <9 days and not varying more than 9 days in length)^100^ and required participants to have at least three consecutive regular cycles prior to vaccination. We also excluded irregular trackers, defined as users who failed to track at least one metric other than menstruation dates, in each of the three cycles preceding vaccination, to improve cycle phase accuracy given known inconsistencies in app-based tracking^101^. Following all other exclusions, one participant with missing ethnicity data was excluded to maintain complete case analysis for multivariable modelling. Participants were classified as vaccinated in the follicular phase (n=760) or luteal phase (n=714) based on their vaccination date within their menstrual cycle. Summary tables with un-collapsed variables in *Sup. Info.* 4.

### Statistical Analysis

All statistical analyses were performed using R (version 4.2.1).

#### Model Specification

To assess whether menstrual cycle phase at the time of vaccination predicted vaccine-related side effects, three separate regression models were used. A binary logistic regression model was used to report presence of side effects, using the *glm* function in R. We modelled reported side effect severity using cumulative odds ordinal logistic regression, implemented using the vglm function from the *VGAM* package with the parallel odds assumption confirmed. The parallel odds assumption was tested using likelihood ratio tests comparing parallel and non-parallel models (p = 0.22), supporting the use of proportional odds model. Number of side effects was modelled with negative binomial regression using the glm.nb function from the *MASS* package, following identification of overdispersion in an initial Poisson model.

To evaluate whether cycle phase predicted subsequent COVID-19 infection outcomes over the follow-up period, we conducted a Wilcoxon rank-sum test to compare time to median infection between the two phases. Time to confirmed infection was then analysed using a Cox proportional hazards model (coxph function from the *survival* package), with time calculated from the date of first vaccination.

Participants were censored at the date of survey completion if they did not report infection. Second vaccine doses were included as time-varying covariates to account for changes in immunity over time.

#### Covariates and model selection

Covariate adjustment sets were selected using directed acyclic graphs (DAG; *Sup. Info.* 2&3). All models testing side effect outcomes were adjusted for age, BMI, ethnicity, smoking status, and presence of pre-existing medical condition(s). Ethnicity was subsequently removed from the final models as its exclusion improved model fit and it did not meaningfully alter results. The Cox proportional hazards model initially included the same variables plus dates of the second and third vaccines, as recommended by the DAG. However, influence diagnostics revealed the full model was unstable due to the small number of COVID-19 cases (n=82) relative to the number of variables. We used stepwise selection to identify the minimum set of variables needed to control for confounding while maintaining model stability. The final Cox model included age, pre-existing conditions, and the second vaccine as a time-varying covariate. Results of the stepwise selection are provided in *Sup. Info*. *12*.

#### Interaction analyses

We tested interactions with cycle phase at vaccination with model covariates age, BMI and smoking but not ethnicity or pre-existing conditions as counts in the “Other” and “Has medical condition” groups were too small and heterogeneous to enable meaningful interpretation. Interactions with Vaccine type, Stress, Copper IUD, Education, Income and Residence were also tested. Only statistically significant results are reported.

#### Sensitivity and exploratory analyses Demographics and health characteristics

We ran several sensitivity analyses. We re-ran our models with complete-case analysis: excluding participants who responded either “Don’t know” to Income or “No answer” to the Stress variable. In categorical variables with a dominant category (>75%) we restricted the sample to that category: “White” (Ethnicity),“More than high school” (Education), “Not fitted” (Copper IUD), “Never” (Smoking), “Average” (Stress) and “mRNA” (Vaccine type). For BMI we re-ran the analysis excluding “Obese”.

#### Menstruation, peri-menstruation and periovulation windows

To evaluate whether concurrent menstrual cycle symptoms disproportionately influenced associations, we conducted three sensitivity analyses excluding participants vaccinated during symptom-heavy windows of the cycle: menstruation, peri-menstruation and periovulation. Menstruation was identified by matching vaccine dates to those falling between ‘period start’ and ‘period end’ dates inclusive in participants’ cycle data. The perimenstrual period was defined as the three days before menses onset through the second day of bleeding (days -3 to +2), capturing this hormonally distinct window^68^. To account for peak estrogen levels and ovulation-specific symptoms, we defined the periovulatory window as days -15 to -12 before the next menstruation (the periovulatory window^59^).

#### Previous symptom tracking

To further evaluate whether such misattribution might have occurred, we used the prospectively collected cycle and tracking data to examine whether participants vaccinated during menstruation, perimenstruation or periovulation had tracked specific symptoms at the same cycle windows in the three cycles prior to their vaccine cycle. We focused on headache and fatigue, as these were among the most reported vaccine side effects and mapped clearly to tracked symptom categories in the app data (’headache’ to “headache“; ‘fatigue’ to “exhausted” and “low energy”). For each participant vaccinated during menstruation, perimenstruation or periovulation, we calculated the number of cycles (0-3) in which they had tracked each symptom during the corresponding cycle window. We then used chi-square tests to assess whether frequency of prior symptom tracking during these cycle windows was associated with reporting that symptom as a vaccine side effect.

#### Alternate cycle-phase classifications

Finally, to assess our cycle phase classification method, we re-ran models with alternative luteal phase lengths. Although 14 days is the classical method for categorising luteal phase length, analysis of 600,000 app-tracked cycles including ovulation, found a mean luteal phase of 12.4 days, with variation by age and cycle length ^61^. We therefore tested luteal phases calculated as: (1) fixed 12 days for all participants, (2) fixed 13 days for all participants, (3) adjusted for age using mean values reported by Bull *et al.* 2019 and (4) adjusted for cycle length using mean values reported by Bull *et al.* 2019. We ran a final model excluding participants who were not classified in the same cycle phase by all these alternative definitions, to assess results in individuals with consistent phase classification across methods.

#### Power

Three side effect analyses (logistic, ordinal, and negative binomial regression) comparing luteal vs follicular phase at vaccination were conducted in the full sample (n = 1,474) and considered jointly for multiple testing. Bonferroni correction was applied to the power calculation and interpretation of primary comparisons, with a corrected significance threshold of α = 0.017. Where relevant, p-values between 0.017 and 0.05 are reported as nominally significant and interpreted with caution. The Cox proportional hazards model for time to COVID-19 infection was treated as a separate hypothesis test with a significance threshold of α = 0.05.

Post hoc power analyses indicated that for logistic and ordinal regression models, assuming Cohen’s f² = 0.02 and 80% power, 872 participants were required. For negative binomial regression assuming a rate ratio of 1.3, 305 participants were required. All three side effect analyses were adequately powered. For the Cox model using Schoenfeld’s method and the observed hazard ratio of 0.73, approximately 320 infection events were required to achieve 80% power. With 82 observed events, the Cox analysis achieved just 29% power.

#### Model Diagnostics and Assumption Checks

Standard diagnostic checks were performed for all models. Multicollinearity was assessed using Variance Inflation Factors, model-specific assumptions were tested (linearity of logit, proportional odds, proportional hazards), and overdispersion was evaluated for count models. Alternate model specifications were evaluated, including models with interaction terms and sensitivity analyses using different cycle phase definitions. Model selection was based on AIC/BIC comparison, with lower values indicating better fit. Key diagnostic results are reported alongside model output.

## Data Availability

Aggregated data are available in Supplementary Information. The underlying database that supports the findings of this study was made available by Clue by BioWink GmbH. While it is de–identified, it cannot be made directly available to the reader. Researchers interested in gaining access to the underlying data can contact Clue by BioWink GmbH and establish a data use agreement with them.

## Supplementary Information

1. Period and the pandemic survey questions
2. DAG – Menstrual Cycle Phase at Vaccination and Side Effects
3. DAG – Menstrual Cycle Phase at Vaccination and COVID-19 Infection
4. Participant characteristics by menstrual cycle phase at first COVID-19 vaccination: un-collapsed variables
5. Association of cycle phase at vaccination with any reported side effects, sub-group and sensitivity analysis results
6. Interaction test results: Cycle phase at vaccination and any reported side effects
7. Association of cycle phase at vaccination with reported severity of side effects, sub-group and sensitivity analysis results
8. Interaction test results: Cycle phase at vaccination and severity of reported side effects
9. Association of cycle phase at vaccination with reported side effect number, sub-group and sensitivity analysis results
10. Interaction test results: Cycle phase at vaccination and number of reported side effects
11. Cox proportional hazards model results table
12. Cox model stepwise selection table
13. Alternate cycle phase estimation model results
14. 18-24 age category and full sample phase comparison

## References

1. Sharp, G. C. et al. The COVID-19 pandemic and the menstrual cycle: research gaps and opportunities. Int. J. Epidemiol. 51, 691–700 (2022).

2. Alvergne, A. Why we must fight ignorance about COVID-19 vaccines and menstrual cycles. Trends Mol. Med. 29, 678–680 (2023).

3. Chao, M. J., Menon, C. & Elgendi, M. Effect of COVID-19 vaccination on the menstrual cycle. Front. Med. 9, 1065421 (2022).

4. Barabás, K. et al. Influence of COVID-19 pandemic and vaccination on the menstrual cycle: A retrospective study in Hungary. Front. Endocrinol. 13, 974788 (2022).

5. Alvergne, A. et al. A retrospective case-control study on menstrual cycle changes following COVID-19 vaccination and disease. iScience 26, 106401 (2023).

6. Alvergne, A., Woon, E. V. & Male, V. Effect of COVID-19 vaccination on the timing and flow of menstrual periods in two cohorts. *Front*. Reprod. Health 4, 952976 (2022).

7. Edelman, A. et al. Association Between Menstrual Cycle Length and Coronavirus Disease 2019 (COVID-19) Vaccination: A U.S. Cohort. Obstet. Gynecol. 10.1097/AOG.0000000000004695 (2022) doi:10.1097/AOG.0000000000004695.

8. Gibson, E. A. et al. Covid-19 vaccination and menstrual cycle length in the Apple Women’s Health Study. Preprint at 10.1101/2022.07.07.22277371 (2022).

9. Ramaiyer, M. et al. The association of COVID-19 vaccination and menstrual health: A period-tracking app-based cohort study. Vaccine X 19, 100501 (2024).

10. Edelman, A. et al. Timing of Coronavirus Disease 2019 (COVID-19) Vaccination and Effects on Menstrual Cycle Changes. Obstet. Gynecol. 10.1097/AOG.0000000000005550 (2024) doi:10.1097/AOG.0000000000005550.

11. Velasco-Regulez, B. et al. Is the phase of the menstrual cycle relevant when getting the covid-19 vaccine? Am. J. Obstet. Gynecol. 227, 913–915 (2022).

12. Sharp, G. C. et al. The COVID-19 pandemic and the menstrual cycle: research gaps and opportunities. Int. J. Epidemiol. 51, 691–700 (2022).

13. Alvergne, A. & Högqvist Tabor, V. Is Female Health Cyclical? Evolutionary Perspectives on Menstruation. Trends Ecol. Evol. 33, 399–414 (2018).

14. Jacobsen, H. & Klein, S. L. Sex Differences in Immunity to Viral Infections. Front. Immunol. 12, 720952 (2021).

15. Klein, S. L. & Flanagan, K. L. Sex differences in immune responses. Nat. Rev. Immunol. 16, 626–638 (2016).

16. Klein, S. L., Jedlicka, A. & Pekosz, A. The Xs and Y of immune responses to viral vaccines. Lancet Infect. Dis. 10, 338–349 (2010).

17. Klein, S. L., Marriott, I. & Fish, E. N. Sex-based differences in immune function and responses to vaccination. Trans. R. Soc. Trop. Med. Hyg. 109, 9–15 (2015).

18. Flanagan, K. L., Fink, A. L., Plebanski, M. & Klein, S. L. Sex and Gender Differences in the Outcomes of Vaccination over the Life Course. Annu. Rev. Cell Dev. Biol. 33, 577–599 (2017).

19. Metcalf, C. J. E. & Graham, A. L. Schedule and magnitude of reproductive investment under immune trade-offs explains sex differences in immunity. Nat. Commun. 9, 4391 (2018).

20. Wilkinson, N. M., Chen, H.-C., Lechner, M. G. & Su, M. A. Sex Differences in Immunity. Annu. Rev. Immunol. 40, 75–94 (2022).

21. Falahi, S. & Kenarkoohi, A. Sex and gender differences in the outcome of patients with COVIDLJ19. J. Med. Virol. 93, 151–152 (2021).

22. Nguyen, N. T. et al. Male gender is a predictor of higher mortality in hospitalized adults with COVID-19. PLOS ONE 16, e0254066 (2021).

23. Scully, E. P., Haverfield, J., Ursin, R. L., Tannenbaum, C. & Klein, S. L. Considering how biological sex impacts immune responses and COVID-19 outcomes. Nat. Rev. Immunol. 20, 442–447 (2020).

24. Nachtigall, I. et al. Effect of gender, age and vaccine on reactogenicity and incapacity to work after COVID-19 vaccination: a survey among health care workers. BMC Infect. Dis. 22, 291 (2022).

25. Rolfes, L. et al. COVID-19 vaccine reactogenicity – A cohort event monitoring study in the Netherlands using patient reported outcomes. Vaccine 40, 970–976 (2022).

26. Sudre, C. H. et al. Attributes and predictors of long COVID. Nat. Med. 27, 626–631 (2021).

27. Bouman, A., Heineman, M. J. & Faas, M. M. Sex hormones and the immune response in humans. Hum. Reprod. Update 11, 411–423 (2005).

28. Ho, J. Q. et al. The immune response to COVID LJ19: Does sex matter? Immunology 166, 429–443 (2022).

29. Pennell, L. M., Galligan, C. L. & Fish, E. N. Sex affects immunity. J. Autoimmun. 38, J282–J291 (2012).

30. Hoffmann, J. P., Liu, J. A., Seddu, K. & Klein, S. L. Sex hormone signaling and regulation of immune function. Immunity 56, 2472–2491 (2023).

31. Roved, J., Westerdahl, H. & Hasselquist, D. Sex differences in immune responses: Hormonal effects, antagonistic selection, and evolutionary consequences. Horm. Behav. 88, 95–105 (2017).

32. Emera, D., Romero, R. & Wagner, G. The evolution of menstruation: A new model for genetic assimilation: Explaining molecular origins of maternal responses to fetal invasiveness. BioEssays 34, 26–35 (2012).

33. Harding, A. T. & Heaton, N. S. The Impact of Estrogens and Their Receptors on Immunity and Inflammation during Infection. Cancers 14, 909 (2022).

34. Newson, L. et al. Sensitive to Infection but Strong in Defense—Female Sex and the Power of Oestradiol in the COVID-19 Pandemic. Front. Glob. Womens Health 2, 651752 (2021).

35. Kaushic, C., Roth, K. L., Anipindi, V. & Xiu, F. Increased prevalence of sexually transmitted viral infections in women: the role of female sex hormones in regulating susceptibility and immune responses. J. Reprod. Immunol. 88, 204–209 (2011).

36. Wira, C. R. & Fahey, J. V. A new strategy to understand how HIV infects women: identification of a window of vulnerability during the menstrual cycle. AIDS 22, 1909–1917 (2008).

37. Chowdhury, N. U., Guntur, V. P., Newcomb, D. C. & Wechsler, M. E. Sex and gender in asthma. Eur. Respir. Rev. 30, 210067 (2021).

38. Colangelo, K., Haig, S., Bonner, A., Zelenietz, C. & Pope, J. Self-reported flaring varies during the menstrual cycle in systemic lupus erythematosus compared with rheumatoid arthritis and fibromyalgia. Rheumatology 50, 703–708 (2011).

39. Latman, N. S. Relation of menstrual cycle phase to symptoms of rheumatoid arthritis. Am. J. Med. 74, 957–960 (1983).

40. Ueno, A., Yoshida, T., Yamamoto, Y. & Hayashi, K. Successful control of menstrual cycleLJrelated exacerbation of inflammatory arthritis with GNRH agonist with addLJback therapy in a patient with rheumatoid arthritis. J. Obstet. Gynaecol. Res. 48, 2005–2009 (2022).

41. Gursoy, A. Y. et al. CRP at early follicular phase of menstrual cycle can cause misinterpretation for cardiovascular risk assessment. Interv. Med. Appl. Sci. 7, 143–146 (2015).

42. Wander, K., Brindle, E. & O’Connor, K. A. CLJreactive protein across the menstrual cycle. Am. J. Phys. Anthropol. 136, 138–146 (2008).

43. Nandi, S. & Rani, R. A study of total leukocyte count in different phases of menstrual cycle. Natl. J. Physiol. Pharm. Pharmacol. 5, 108 (2015).

44. Lee, S. et al. Fluctuation of Peripheral Blood T, B, and NK Cells during a Menstrual Cycle of Normal Healthy Women. J. Immunol. 185, 756–762 (2010).

45. Okimura, H. et al. Changes in the proportion of regulatory T cell subpopulations during menstrual cycle and early pregnancy. Am. J. Reprod. Immunol. 88, e13636 (2022).

46. Whettlock, E. M. et al. Dynamic Changes in Uterine NK Cell Subset Frequency and Function Over the Menstrual Cycle and Pregnancy. Front. Immunol. 13, 880438 (2022).

47. Dunn, S. E., Perry, W. A. & Klein, S. L. Mechanisms and consequences of sex differences in immune responses. Nat. Rev. Nephrol. 20, 37–55 (2024).

48. Klein, S. L. & Morgan, R. The impact of sex and gender on immunotherapy outcomes. Biol. Sex Differ. 11, 24 (2020).

49. Vom Steeg, L. G. & Klein, S. L. Sex and sex steroids impact influenza pathogenesis across the life course. Semin. Immunopathol. 41, 189–194 (2019).

50. Anesi, N., Miquel, C.-H., Laffont, S. & Guéry, J.-C. The Influence of Sex Hormones and X Chromosome in Immune Responses. in Sex and Gender Differences in Infection and Treatments for Infectious Diseases (eds Klein, S. L. & Roberts, C. W.) vol. 441 21–59 (Springer International Publishing, Cham, 2023).

51. Bernstein, S. R., Kelleher, C. & Khalil, R. A. Gender-based research underscores sex differences in biological processes, clinical disorders and pharmacological interventions. Biochem. Pharmacol. 215, 115737 (2023).

52. Alvergne, A. et al. Associations Among Menstrual Cycle Length, Coronavirus Disease 2019 (COVID-19), and Vaccination. Obstet. Gynecol. 143, 83–91 (2024).

53. Li, H. et al. Menstrual cycle length variation by demographic characteristics from the Apple Women’s Health Study. Npj Digit. Med. 6, 100 (2023).

54. Hannan, K. et al. Mood symptoms and gut function across the menstrual cycle in individuals with premenstrual syndrome. Horm. Behav. 166, 105634 (2024).

55. Shea, A. A., Wever, F., Ventola, C., Thornburg, J. & Vitzthum, V. J. More than blood: app-tracking reveals variability in heavy menstrual bleeding construct. BMC Womens Health 23, 170 (2023).

56. deSouza, P. N., et al. The effect of air pollution exposure on menstrual cycle health using self-reported data from a mobile health app: a prospective, observational study. *Lancet Planet*. Health 9, e364–e373 (2025).

57. Zaidi, S. et al. COVID-19 vaccines side effects among the general population during the pandemic: a cross-sectional study. Front. Public Health 13, 1420291 (2025).

58. CDC. Coronavirus Disease 2019 (COVID-19) Vaccine Safety.

59. Schmalenberger, K. M. et al. How to study the menstrual cycle: Practical tools and recommendations. Psychoneuroendocrinology 123, 104895 (2021).

60. Callahan, D. Managed Care and the Goals of Medicine. J. Am. Geriatr. Soc. 46, 385–388 (1998).

61. Bull, J. R. et al. Real-world menstrual cycle characteristics of more than 600,000 menstrual cycles. Npj Digit. Med. 2, 83 (2019).

62. UNICEF. Menstrual hygiene. https://www.unicef.org/wash/menstrual-hygiene (n.d.).

63. Liu, K. A. & DiPietro Mager, N. A. Women’s involvement in clinical trials: historical perspective and future implications. Pharm. Pract. 14, 708–708 (2016).

64. Zuk, M. The Sicker Sex. PLoS Pathog. 5, e1000267 (2009).

65. Mason, L. et al. ‘We Keep It Secret So No One Should Know’ – A Qualitative Study to Explore Young Schoolgirls Attitudes and Experiences with Menstruation in Rural Western Kenya. PLoS ONE 8, e79132 (2013).

66. Tingen, C. M., Halvorson, L. M. & Bianchi, D. W. Revisiting menstruation: the misery, mystery, and marvel. Am. J. Obstet. Gynecol. 223, 617–618 (2020).

67. Critchley, H. O. D. et al. Menstruation: science and society. Am. J. Obstet. Gynecol. 223, 624–664 (2020).

68. Schmalenberger, K. M. et al. How to study the menstrual cycle: Practical tools and recommendations. Psychoneuroendocrinology 123, 104895 (2021).

69. Lorenz, T. K., Worthman, C. M. & Vitzthum, V. J. Links among inflammation, sexual activity and ovulation: Evolutionary trade-offs and clinical implications. Evol. Med. Public Health 2015, 304–324 (2015).

70. Case, A. M. & Reid, R. L. Effects of the Menstrual Cycle on Medical Disorders. Arch. Intern. Med. 158, 1405 (1998).

71. Khan, D. & Ansar Ahmed, S. The Immune System Is a Natural Target for Estrogen Action: Opposing Effects of Estrogen in Two Prototypical Autoimmune Diseases. Front. Immunol. 6, (2016).

72. Zwahlen, M. & Stute, P. Impact of progesterone on the immune system in women: a systematic literature review. Arch. Gynecol. Obstet. 309, 37–46 (2023).

73. Moulton, V. R. Sex Hormones in Acquired Immunity and Autoimmune Disease. Front. Immunol. 9, 2279 (2018).

74. Hughes, G. C. Progesterone and autoimmune disease. Autoimmun. Rev. 11, A502–A514 (2012).

75. Dilbaz, B. & Aksan, A. Premenstrual syndrome, a common but underrated entity: review of the clinical literature. J. Turk.-Ger. Gynecol. Assoc. 22, 139–148 (2021).

76. Epperson, C. N. et al. Premenstrual Dysphoric Disorder: Evidence for a New Category for DSM-5. Am. J. Psychiatry 169, 465–475 (2012).

77. Granella, F. et al. Migraine Without Aura and Reproductive Life Events: A Clinical Epidemiological Study in 1300 Women. Headache J. Head Face Pain 33, 385–389 (1993).

78. Brott, N., R. & Le, J., K. Mittelschmerz. in StatPearls (StatPearls Publishing, Treasure Island (FL), 2023).

79. Shen, H.-H. et al. Ovarian hormones-autophagy-immunity axis in menstruation and endometriosis. Theranostics 11, 3512–3526 (2021).

80. Brabaharan, S. et al. Association of Hormonal Contraceptive Use With Adverse Health Outcomes: An Umbrella Review of Meta-analyses of Randomized Clinical Trials and Cohort Studies. *JAMA Netw*. Open 5, e2143730 (2022).

81. Shaikh, S. R., Beck, M. A., Alwarawrah, Y. & MacIver, N. J. Emerging mechanisms of obesity-associated immune dysfunction. Nat. Rev. Endocrinol. 20, 136–148 (2024).

82. Monteiro, R., Teixeira, D. & Calhau, C. Estrogen Signaling in Metabolic Inflammation. Mediators Inflamm. 2014, 1–20 (2014).

83. Santoro, N., Roeca, C., Peters, B. A. & Neal-Perry, G. The Menopause Transition: Signs, Symptoms, and Management Options. J. Clin. Endocrinol. Metab. 106, 1–15 (2021).

84. Radzey, N. et al. Genital inflammatory status and the innate immune response to contraceptive initiation. Am. J. Reprod. Immunol. 88, e13542 (2022).

85. Chou, C.-H. et al. Divergent endometrial inflammatory cytokine expression at peri-implantation period and after the stimulation by copper intrauterine device. Sci. Rep. 5, 15157 (2015).

86. Florek, E. et al. Differences in the sex hormone levels in the menstrual cycle due to tobacco smoking - a myth or reality? Endokrynol. Pol. VM/OJS/J/85458 (2021) doi:10.5603/EP.a2021.0097.

87. Rosen Vollmar, A. K., Mahalingaiah, S. & Jukic, A. M. The menstrual cycle as a vital sign: a comprehensive review. FS Rev. 6, 100081 (2025).

88. Ponticelli, D. et al. Smoking habits predict adverse effects after mRNA COVID-19 vaccine: Empirical evidence from a pilot study. Public Health 219, 18–21 (2023).

89. Grieger, J. A. & Norman, R. J. Menstrual Cycle Length and Patterns in a Global Cohort of Women Using a Mobile Phone App: Retrospective Cohort Study. J. Med. Internet Res. 22, e17109 (2020).

90. George, K. & Kamath, M. S. Fertility and age. J. Hum. Reprod. Sci. 3, 121–123 (2010).

91. Wahl, I. & Wardemann, H. Sterilizing immunity: Understanding COVID-19. Immunity 55, 2231–2235 (2022).

92. Beukenhorst, A. L. et al. SARS-CoV-2 elicits non-sterilizing immunity and evades vaccine-induced immunity: implications for future vaccination strategies. Eur. J. Epidemiol. 38, 237–242 (2023).

93. Hermann, E. A. et al. Association of Symptoms After COVID-19 Vaccination With Anti–SARS-CoV-2 Antibody Response in the Framingham Heart Study. *JAMA Netw*. Open 5, e2237908 (2022).

94. Jorda, A. et al. Association between reactogenicity and immunogenicity after BNT162b2 booster vaccination: a secondary analysis of a prospective cohort study. Clin. Microbiol. Infect. Off. Publ. Eur. Soc. Clin. Microbiol. Infect. Dis. 29, 1188–1195 (2023).

95. Sutton, D. et al. COVID-19 vaccine acceptance among pregnant, breastfeeding, and nonpregnant reproductive-aged women. Am. J. Obstet. Gynecol. MFM 3, 100403 (2021).

96. Robertson, E. et al. Predictors of COVID-19 vaccine hesitancy in the UK household longitudinal study. Brain. Behav. Immun. 94, 41–50 (2021).

97. Gupta, S., Rai, D. & Shukla, S. COVID vaccine: Social, menstrual and psychological aftermath. Vacunas Engl. Ed. 24, 326–334 (2023).

98. Weir, C. B. & Jan, A. BMI Classification Percentile And Cut Off Points. in StatPearls (StatPearls Publishing, Treasure Island (FL), 2025).

99. Beladiya, J. et al. Safety and efficacy of COVIDLJ19 vaccines: A systematic review and metaLJanalysis of controlled and randomized clinical trials. Rev. Med. Virol. 34, e2507 (2024).

100. Munro, M. G. et al. The FIGO ovulatory disorders classification system. Int. J. Gynecol. Obstet. 159, 1–20 (2022).

101. Schantz, J. S., Fernandez, C. S. P. & Jukic, A. M. Z. Menstrual Cycle Tracking Applications and the Potential for Epidemiological Research: a Comprehensive Review of the Literature. Curr. Epidemiol. Rep. 8, 9–19 (2021).

