## Supplementary Information 1 for "Menstrual cycle phase and its association with COVID-19 vaccine side effects and subsequent infection: A study of period tracking app users"

### Periods and the pandemic

---

1. **Please confirm that you have read the information above and agree with the following statements:** *

- ☐ I have read the information and voluntarily agree to participate in this study
- ☐ I am 18 years of age or older
- ☐ I experience menstrual periods (or withdrawal bleeds)
- ☐ I have not been pregnant or breastfeeding since January 2020

2. **How old are you?**

Clue_ID **Action: Hidden Value**
Value: [url("id")]

**(untitled)**

**Page exit logic:** Skip / Disqualify Logic
**IF:** #3 Question "

**Where do you currently live?**

" is one of the following answers ("Other") **THEN:** Disqualify and display:
Thank you for supporting Clue research. For this survey, we're focusing on just a few countries, but we will have more surveys for you in the future!

LOGIC Show/hide trigger exists.

3. **Where do you currently live?**

- United States of America
- United Kingdom
- Australia
- Canada
- Other

**(untitled)**

---

**Page exit logic:** Disqualify respondents from other countries
**IF:** #3 Question "

**Where do you currently live?**

" is not one of the following answers ("United States of America","United Kingdom","Australia","Canada")**THEN:** Disqualify and display:
This specific study focuses on people living in Australia, Canada, the United States of America and the United Kingdom. Thank you for your interest in contributing to our research!

LOGIC Hidden unless: #3 Question "

**Where do you currently live?**

" is one of the following answers ("United Kingdom")

4. **Which country do you live in?**

- England
- Scotland
- Wales
- Northern Ireland
- Other - Write In

Hidden unless: #3 Question "
**Where do you currently live?**

" is one of the following answers ("United States of America")

5. **Which state do you live in?**

- Alabama
- Alaska
- Arizona
- Arkansas
- California
- Colorado
- Connecticut
- Delaware
- Florida
- Georgia
- Hawaii
- Idaho
- Illinois
- Indiana
- Iowa
- Kansas
- Kentucky
- Louisiana
- Maine
- Maryland
- Massachusetts
- Michigan
- Minnesota
- Mississippi
- Missouri
- Montana
- Nebraska
- Nevada
- New Hampshire
- New Jersey
- New Mexico
- New York
- North Carolina
- North Dakota
- Ohio
- Oklahoma
- Oregon
- Pennsylvania
- Puerto Rico
- Rhode Island
- South Carolina
- South Dakota
- Tennessee
- Texas
- Utah
- Vermont
- Virginia
- Washington
- Washington, D.C.
- West Virginia
- Wisconsin
- Wyoming
- Other US territory

LOGIC Hidden unless: #3 Question "
**Where do you currently live?**

" is one of the following answers ("Canada")

6. **Which province do you live in?**

- Alberta
- British Columbia
- Manitoba
- New Brunswick
- Newfoundland and Labrador
- Nova Scotia
- Ontario
- Prince Edward Island
- Quebec
- Saskatchewan
- Other - Write In

LOGIC Hidden unless: #3 Question "
**Where do you currently live?**

" is one of the following answers ("Australia")

7. **Which state or territory do you live in?**

- Northern Territory
- New South Wales
- Australian Capital Territory
- Queensland
- South Australia
- Tasmania
- Victoria
- Western Australia
- External Territories
- Other - Write In

LOGIC Show/hide trigger exists.

8. **We need to ask for your weight - do you prefer kilograms (kg), pounds (lbs) or stones?**

- kg
- lbs
- stones

LOGIC Hidden unless: #8 Question "**We need to ask for your weight - do you prefer kilograms (kg), pounds (lbs) or stones?** is one of the following answers ("kg")

9. **How much do you weigh in kg?**

[illegible]

120+
I don't know
Prefer not to say

LOGIC Hidden unless: #8 Question "**We need to ask for your weight - do you prefer kilograms (kg), pounds (lbs) or stones?** is one of the following answers ("lbs")

10. **How much do you weigh in pounds (lbs)?**

[illegible]

265+
I don't know
Prefer not to say

LOGIC Hidden unless: #8 Question "**We need to ask for your weight - do you prefer kilograms (kg), pounds (lbs) or stones?**" is one of the following answers ("stones")

11. **How much do you weigh in stones?**

6s
6s1
6s2
6s3
6s4
6s5
6s6
6s7
6s8
6s9
6s10
6s11
6s12
6s13
7s
7s1
7s2
7s3
7s4
7s5
7s6
7s7
7s8
7s9
7s10
7s11
7s12
7s13
8s
8s1
8s2
8s3
8s4
8s5
8s6

8s7
8s8
8s9
8s10
8s11
8s12
8s13
9s
9s1
9s2
9s3
9s4
9s5
9s6
9s7
9s8
9s9
9s10
9s11
9s12
9s13
10s
10s1
10s2
10s3
10s4
10s5
10s6
10s7
10s8
10s9
10s10
10s11
10s12
10s13
11s
11s1
11s2
11s3
11s4
11s5
11s6
11s7
11s8
11s9
11s10
11s11
11s12
11s13
12s
12s1
12s2
12s3
12s4
12s5
12s6
12s7
12s8
12s9
12s10
12s11
12s12
12s13
13s
13s1
13s2
13s3
13s4
13s5
13s6
13s7
13s8
13s9
[illegible]

13s10
13s11
13s12
13s13
14s
14s1
14s2
14s3
14s4
14s5
14s6
14s7
14s8
14s9
14s10
14s11
14s12
14s13
15s
15s1
15s2
15s3
15s4
15s5
15s6
15s7
15s8
15s9
15s10
15s11
15s12
15s13
16s
16s1
16s2
16s3
16s4
16s5
16s6
16s7
16s8
16s9
16s10
16s11
16s12
16s13
17s
17s1
17s2
17s3
17s4
17s5
17s6
17s7
17s8
17s9
17s10
17s11
17s12
17s13
18s
18s1
18s2
18s3
18s4
18s5
18s6
18s7
18s8
18s9
18s10
18s11
18s12
18s13

[illegible]
19s
19s1
19s2
19s3
19s4
19s5
19s6
19s7
19s8
19s9
19s10
19s11
19s12
19s13
20s
20s1
20s2
20s3
20s4
20s5
20s6
20s7
20s8
20s9
20s10
20s11
20s12
20s13
I don't know
Prefer not to say

LOGIC Show/hide trigger exists.

12. **We need to ask for your height - do you prefer centimetres (cm) or feet & inches?**

- cm
- feet & inches

LOGIC Hidden unless: #12 Question "**We need to ask for your height - do you prefer centimetres (cm) or feet & inches?**" is one of the following answers ("cm")

13. **What is your height in cm?**

[illegible]

>210

LOGIC Hidden unless: #12 Question "**We need to ask for your height - do you prefer centimetres (cm) or feet & inches?**" is one of the following answers ("feet & inches")

14. **What is your height in feet and inches?**

4' 0
4' 1"
4' 2
4' 3
4' 4
4' 5
4' 6
4' 7
4' 8
4' 9
4' 10
4' 11
5' 0
5' 1"
5' 2
5' 3
5' 4
5' 5
5' 6
5' 7
5' 8
5' 9
5' 10
5' 11
6' 0
6' 1"
6' 2
6' 3
6' 4
6' 5
6' 6
6' 7
6' 8
6' 9
6' 10
6' 11

15. **What is the highest level of education you have completed?**

- Less than high school
- Some high school
- High school diploma (A-levels, BTEC, AP)
- Some college or University
- Undergraduate degree (e.g. Bachelor's degree)
- Post-graduate degree (e.g. Master's, PhD, MD, JD)

16. **How many children have you given birth to?**

- 0
- 1
- 2
- 3
- 4
- 5+

LOGIC Show/hide trigger exists.

17. **Have you ever smoked tobacco or vaped e-cigarettes?** Select those that you have ever done

- Smoked tobacco
- Vaped e-cigarettes
- None

LOGIC Hidden unless: #17 Question "**Have you ever smoked tobacco or vaped e-cigarettes?**Select those that you have ever done" is one of the following answers ("Smoked tobacco")

18. **How often do you smoke tobacco?**

- I have not smoked in the past 2 years
- Several times a day
- Once a day
- Once a week
- Once a month
- Less than a few times per year

LOGIC Hidden unless: #17 Question "**Have you ever smoked tobacco or vaped e-cigarettes?**Select those that you have ever done" is one of the following answers ("Vaped e-cigarettes")

19. **How often do you vape e-cigarettes?**

- I have not vaped in the past 2 years
- Several times a day
- Once a day
- Once a week
- Once a month
- Less than a few times per year

LOGIC Show/hide trigger exists.

20. **Have you ever smoked cannabis?**

- No
- Yes

LOGIC Hidden unless: #20 Question "**Have you ever smoked cannabis?**" is one of the following answers ("Yes")

21. **How often do you smoke cannabis?**

- I do not currently smoke cannabis
- Several times a day
- Once a day
- Once a week
- Once a month
- Less than a few times a year

22. **What do you identify as?**

- Woman
- Man
- Genderqueer / Non-Binary
- Questioning/ Unsure
- Prefer not to answer
- Other (Please specify)

LOGIC Hidden unless: #3 Question "
**Where do you currently live?**

" is one of the following answers ("United States of America")

23. **How would you describe your ethnic identity?** (check all that apply)

- ☐ White or European American
- ☐ Hispanic, Latina or Latinx
- ☐ Black or African-American
- ☐ Asian or Asian-American
- ☐ Native American or Alaska native
- ☐ Native Hawaiian or other Pacific Islander
- ☐ Middle Eastern / North African
- ☐ Prefer not to say

LOGIC Hidden unless: #3 Question "
**Where do you currently live?**

" is one of the following answers ("United Kingdom")

24. **How would you describe your ethnic identity?** (check all that apply)

- ☐ White European
- ☐ Black British or Afro-Caribbean
- ☐ Asian or Asian British
- ☐ Middle Eastern or Middle Eastern British
- ☐ Roma/Gypsy/Traveller/Irish Traveller
- ☐ Prefer not to say

LOGIC Hidden unless: #3 Question "
**Where do you currently live?**

" is one of the following answers ("Canada")

25. **How would you describe your ethnic identity?** (check all that apply)

- ☐ First Nations, Indigenous, Aboriginal, or Native peoples
- ☐ Northern Asian (Chinese, Japanese, Korean)
- ☐ Southern Asian (India, Pakistan)
- ☐ Southeast Asian & Filipino
- ☐ Black Canadian, Caribbean Canadian, or African Canadian
- ☐ White (European ancestry)
- ☐ Arab, Middle Eastern, or North African
- ☐ Latin American
- ☐ Prefer not to say

LOGIC Hidden unless: #3 Question "
**Where do you currently live?**

" is one of the following answers ("Australia")

26. **How would you describe your ethnic identity?** (check all that apply)

- ☐ White
- ☐ Asian
- ☐ Black or African
- ☐ Aboriginal
- ☐ Middle Eastern
- ☐ Pacific Islander
- ☐ Prefer not to say

Hidden unless: #3 Question "
**Where do you currently live?**

" is one of the following answers ("United Kingdom")

27. **What is the total take-home pay** *(the money you have after all taxes and contributions have been deducted, also known as Net Income)* **earned by you and other people in your household (your partner/family)?** *If you live on campus or with your parents, and if you rely on your parents' income more than you rely on your own income, then indicate your parents' income.*

- Less than £13,680 (year) OR £1,140 (month)
- £13,680 to £22,140 (year) OR £1,140 to £1,840 (month)
- £22,140 to £29,250 (year) OR £1,840 to £2,440 (month)
- £29,250 to £39,400 (year) OR £2,440 to £3,280 (month)
- £39,400 to £76,140 (year) OR £3,280 to £6,340 (month)
- More than £76,140 (year) OR £6,340 (month)
- Don't know

Hidden unless: #3 Question "
**Where do you currently live?**

" is one of the following answers ("United States of America")

28. **What is the total take-home pay** *(the money you have after all taxes and contributions have been deducted, also known as Net Income)* **earned by you and other people in your household (your partner/family)?** *If you live on campus or with your parents, and if you rely on your parents' income more than you rely on your own income, then indicate your parents' income.*

- Less than $13,260 (year) OR $1,100 (month)
- $13,260 to $35,400 (year) OR $1,100 to $2,950 (month)
- $35,400 to $61,560 (year) OR $2,950 to $5,130 (month)
- $61,560 to $99,030 (year) OR $5,130 to $8,250 (month)
- $99,030 to $221,850 (year)OR $8,250 to $18,490 (month)
- More than $221,850 (year) OR $18,490 (month)
- Don't know

Hidden unless: #3 Question "
**Where do you currently live?**

" is one of the following answers ("Canada")

29. **What is the total take-home pay** *(the money you have after all taxes and contributions have been deducted, also known as Net Income)* **earned by you and other people in your household (your partner/family)?** *If you live on campus or with your parents, and if you rely on your parents' income more than you rely on your own income, then indicate your parents' income.*

- Less than CA$25,430 (year) OR CA$2,119 (month)
- CA$25,430 to CA$52,010 (year) OR CA$2,119 to CA$4,330 (month)
- CA$52,010 to CA$71,150 (year) OR CA$4,330 to CA$5,930 (month)
- CA$71,150 to CA$95,200 (year) OR CA$5,930 to CA$7,930 (month)
- CA$95,200 to CA$162,960 (year) OR CA$7,930 to CA$13,580 (month)
- More than CA$162,960 (year) OR CA$13,580 (month)
- Don't know

Hidden unless: #3 Question "
**Where do you currently live?**

" is one of the following answers ("Australia")

30. **What is the total take-home pay** *(the money you have after all taxes and contributions have been deducted, also known as Net Income)* **earned by you and other people in your household (your partner/family)?** *If you live on campus or with your parents, and if you rely on your parents' income more than you rely on your own income, then indicate your parents' income.*

- Less than $13,960 (year) OR $1,160 (month)
- $13,960 to $40,400 (year) OR $1,160 to $3,370 (month)
- $40,400 to $79,780 (year) OR $3,370 to $6,650 (month)
- $79,780 to $123,490 (year) OR $6,650 to $10,290(month)
- $123,490 to $218,840 (year) OR $10,290 to $18,240 (month)
- More than $218,840 (year) OR $18,240 (month)
- Don't know

31. **Have you been diagnosed with any of the following conditions?** (check all that apply)

- ☐ Endometriosis
- ☐ Polycystic ovary syndrome (PCOS)
- ☐ Thyroid disease
- ☐ Uterine polyps or fibroids
- ☐ Eating disorder (e.g. anorexia, bulimia, binge-eating...)
- ☐ Pelvic inflammatory disease
- ☐ Interstitial cystitis
- ☐ Diabetes
- ☐ Gynecological cancers
- ☐ Any other cancer
- ☐ Von Willebrand disease
- ☐ Abnormal pap smear
- ☐ Condition requiring immunosuppresive therapy
- ☐ None of the above
- ☐ Prefer not to answer

32. **How many people can you count on if you have significant personal problems?** This can include family, friends, coworkers, neighbors, etc.

- ○ None
- ○ 1–2
- ○ 3–5
- ○ 5+

33. **How easy is it to get practical help from neighbors** (e.g. someone who lives close to you but is not necessarily family/friends) **in case of urgent need?** For example, if you need to borrow money or a car, or if you need help coping with a crisis.

- ○ Very difficult
- ○ Difficult
- ○ Possible
- ○ Easy
- ○ Very easy

34. **How much interest and concern do people show in what you do?** *This includes your work, interests and/or hobbies.*

- None
- Little
- Uncertain
- Some
- A lot

---

Show/hide trigger exists.

35. **Were you using a hormonal contraceptive or a copper IUD in January 2020?**

- Yes
- No

Show/hide trigger exists. Hidden unless: #35 Question "**Were you using a hormonal contraceptive or a copper IUDin January 2020?** " is one of the following answers ("Yes")

36. **Select all the method(s) you were using in January 2020.**

- Combined oral contraceptive pill (e.g. Microgynon, Marvelon, Yasmin)
- Progestin only pill (e.g. Minipill)
- Contraceptive Injection (e.g. Depo-Provera, Sayana Press, Noristerat)
- Hormonal IUD/IUS/Coil (e.g. Mirena, Kyleena, Jaydess, Levosert or Skyla)
- Contraceptive patch (e.g. Ortho Evra, Xulane
- Vaginal ring (e.g. Nuvaring)
- Contraceptive Implant (e.g. Nexplanon)
- Copper IUD
- None of the above

Show/hide trigger exists. Hidden unless: #36 Question "
**Select all the method(s) you were using in January 2020.**

" is one of the following answers ("Combined oral contraceptive pill (e.g. Microgynon, Marvelon, Yasmin)")

37. **Did you use the combined oral contraceptive pill for the entire pandemic (since 2020)?**

- Yes
- No

Show/hide trigger exists. Hidden unless: #36 Question "
**Select all the method(s) you were using in January 2020.**

" is one of the following answers ("Progestin only pill (e.g. Minipill)")

38. **Did you use the progestin only pill for the entire pandemic (since 2020)?**

- Yes
- No

Show/hide trigger exists. Hidden unless: #36 Question "
**Select all the method(s) you were using in January 2020.**

" is one of the following answers ("Contraceptive Injection (e.g. Depo-Provera, Sayana Press, Noristerat)")

39. **Did you use the contraceptive injection for the entire pandemic (since 2020)?**

- Yes
- No

Show/hide trigger exists. Hidden unless: #36 Question "
**Select all the method(s) you were using in January 2020.**

" is one of the following answers ("Hormonal IUD/IUS/Coil (e.g. Mirena, Kyleena, Jaydess, Levosert or Skyla)")

40. **Did you use the hormonal IUD/IUS/Coil for the entire pandemic (since 2020)?**

- Yes
- No

Show/hide trigger exists. Hidden unless: #36 Question "
**Select all the method(s) you were using in January 2020.**

" is one of the following answers ("Contraceptive patch (e.g. Ortho Evra, Xulane")

41. **Did you use the contraceptive patch for the entire pandemic (since 2020)?**

- Yes
- No

Show/hide trigger exists. Hidden unless: #36 Question "
**Select all the method(s) you were using in January 2020.**

" is one of the following answers ("Vaginal ring (e.g. Nuvaring)")

42. **Did you use the vaginal ring for the entire pandemic (since 2020)?**

- Yes
- No

LOGIC Show/hide trigger exists. Hidden unless: #36 Question "
**Select all the method(s) you were using in January 2020.**

" is one of the following answers ("Contraceptive Implant (e.g. Nexplanon)")

43. **Did you use the contraceptive implant for the entire pandemic (since 2020)?**

- ○ Yes
- ○ No

LOGIC Show/hide trigger exists. Hidden unless: #36 Question "
**Select all the method(s) you were using in January 2020.**

" is one of the following answers ("Copper IUD")

44. **Did you use the copper IUD for the entire pandemic (since 2020)?**

- ○ Yes
- ○ No

LOGIC Hidden unless: #37 Question "
**Did you use the combined oral contraceptive pill for the entire pandemic (since 2020)?**

" is one of the following answers ("No")

45. **When did you stop using the combined oral contraceptive pill?** *If you used it for more than one time period, when did you stop the first time?*

Jan 2020
Feb 2020
March 2020
April 2020
May 2020
June 2020
July 2020
August 2020
Sept 2020
Oct 2020
Nov 2020
Dec 2020
Jan 2021
Feb 2021
March 2021
April 2021
May 2021
June 2021
July 2021
August 2021
Sept 2021
Oct 2021
Nov 2021
Dec 2021

LOGIC Hidden unless: #38 Question "
**Did you use the progestin only pill for the entire pandemic (since 2020)?**

" is one of the following answers ("No")

46. **When did you stop using the progesterone only pill?** *If you used it for more than one time period, when did you stop the first time?*

- Jan 2020
- Feb 2020
- March 2020
- April 2020
- May 2020
- June 2020
- July 2020
- August 2020
- Sept 2020
- Oct 2020
- Nov 2020
- Dec 2020
- Jan 2021
- Feb 2021
- March 2021
- April 2021
- May 2021
- June 2021
- July 2021
- August 2021
- Sept 2021
- Oct 2021
- Nov 2021
- Dec 2021

LOGIC Hidden unless: #39 Question "
**Did you use the contraceptive injection for the entire pandemic (since 2020)?**

" is one of the following answers ("No")

47. **When did you stop using the contraceptive injection?** *If you used it for more than one time period, when did you stop the first time?*

- Jan 2020
- Feb 2020
- March 2020
- April 2020
- May 2020
- June 2020
- July 2020
- August 2020
- Sept 2020
- Oct 2020
- Nov 2020
- Dec 2020
- Jan 2021
- Feb 2021
- March 2021
- April 2021
- May 2021
- June 2021
- July 2021
- August 2021
- Sept 2021
- Oct 2021
- Nov 2021
- Dec 2021

LOGIC Hidden unless: #40 Question "
**Did you use the hormonal IUD/IUS/Coil for the entire pandemic (since 2020)?**

" is one of the following answers ("No")

48. **When did you stop using the hormonal IUD/IUS/Coil?** *If you used it for more than one time period, when did you stop the first time?*

- Jan 2020
- Feb 2020
- March 2020
- April 2020
- May 2020
- June 2020
- July 2020
- August 2020
- Sept 2020
- Oct 2020
- Nov 2020
- Dec 2020
- Jan 2021
- Feb 2021
- March 2021
- April 2021
- May 2021
- June 2021
- July 2021
- August 2021
- Sept 2021
- Oct 2021
- Nov 2021
- Dec 2021

LOGIC Hidden unless: #41 Question "
**Did you use the contraceptive patch for the entire pandemic (since 2020)?**

" is one of the following answers ("No")

49. **When did you stop using the contraceptive patch?** *If you used it for more than one time period, when did you stop the first time?*

- Jan 2020
- Feb 2020
- March 2020
- April 2020
- May 2020
- June 2020
- July 2020
- August 2020
- Sept 2020
- Oct 2020
- Nov 2020
- Dec 2020
- Jan 2021
- Feb 2021
- March 2021
- April 2021
- May 2021
- June 2021
- July 2021
- August 2021
- Sept 2021
- Oct 2021
- Nov 2021
- Dec 2021

LOGIC Hidden unless: #43 Question "
**Did you use the contraceptive implant for the entire pandemic (since 2020)?**

" is one of the following answers ("No")

50. **When did you stop using the contraceptive implant?** *If you used it for more than one time period, when did you stop the first time?*

Jan 2020
Feb 2020
March 2020
April 2020
May 2020
June 2020
July 2020
August 2020
Sept 2020
Oct 2020
Nov 2020
Dec 2020
Jan 2021
Feb 2021
March 2021
April 2021
May 2021
June 2021
July 2021
August 2021
Sept 2021
Oct 2021
Nov 2021
Dec 2021

LOGIC Hidden unless: #44 Question "
**Did you use the copper IUD for the entire pandemic (since 2020)?**

" is one of the following answers ("No")

51. **When did you stop using the copper IUD?** *If you used it for more than one time period, when did you stop the first time?*

Jan 2020
Feb 2020
March 2020
April 2020
May 2020
June 2020
July 2020
August 2020
Sept 2020
Oct 2020
Nov 2020
Dec 2020
Jan 2021
Feb 2021
March 2021
April 2021
May 2021
June 2021
July 2021
August 2021
Sept 2021
Oct 2021
Nov 2021
Dec 2021

LOGIC Show/hide trigger exists. Hidden unless: #37 Question "
**Did you use the combined oral contraceptive pill for the entire pandemic (since 2020)?**

" is one of the following answers ("No")

52. **Did you use any hormonal method or copper IUD after you stopped using combined oral contraceptives? Select which method you used next.** *If you started the same method again, select that method.*

- Combined oral contraceptive pill (e.g. Microgynon, Marvelon, Yasmin)
- Progestin only pill (e.g. Minipill)
- Contraceptive Injection (e.g. Depo-Provera, Sayana Press, Noristerat)
- Hormonal IUD/IUS/Coil (e.g. Mirena, Kyleena, Jaydess, Levosert or Skyla)
- Contraceptive patch (e.g. Ortho Evra, Xulane
- Vaginal ring (e.g. Nuvaring)
- Contraceptive Implant (e.g. Nexplanon)
- Copper IUD
- None of the above

LOGIC Show/hide trigger exists. Hidden unless: #38 Question "
**Did you use the progestin only pill for the entire pandemic (since 2020)?**

" is one of the following answers ("No")

53. **Did you use any hormonal method or copper IUD after you stopped using the progestin-only pill?** *Select which method you used next. If you started the same method again, select that method.*

- Combined oral contraceptive pill (e.g. Microgynon, Marvelon, Yasmin)
- Progestin only pill (e.g. Minipill)
- Contraceptive Injection (e.g. Depo-Provera, Sayana Press, Noristerat)
- Hormonal IUD/IUS/Coil (e.g. Mirena, Kyleena, Jaydess, Levosert or Skyla)
- Contraceptive patch (e.g. Ortho Evra, Xulane
- Vaginal ring (e.g. Nuvaring)
- Contraceptive Implant (e.g. Nexplanon)
- Copper IUD
- None of the above

LOGIC Show/hide trigger exists. Hidden unless: #39 Question "
**Did you use the contraceptive injection for the entire pandemic (since 2020)?**

" is one of the following answers ("No")

54. **Did you use any hormonal method or copper IUD after you stopped using the contraceptive injection?** *Select which method you used next. If you started the same method again, select that method.*

- Combined oral contraceptive pill (e.g. Microgynon, Marvelon, Yasmin)
- Progestin only pill (e.g. Minipill)
- Contraceptive Injection (e.g. Depo-Provera, Sayana Press, Noristerat)
- Hormonal IUD/IUS/Coil (e.g. Mirena, Kyleena, Jaydess, Levosert or Skyla)
- Contraceptive patch (e.g. Ortho Evra, Xulane
- Vaginal ring (e.g. Nuvaring)
- Contraceptive Implant (e.g. Nexplanon)
- Copper IUD
- None of the above

LOGIC Show/hide trigger exists. Hidden unless: #40 Question "
**Did you use the hormonal IUD/IUS/Coil for the entire pandemic (since 2020)?**

" is one of the following answers ("No")

55. **Did you use any hormonal method or copper IUD after you stopped using the hormonal IUD/IUS/Coil?** *Select which method you used next. If you started the same method again, select that method.*

- Combined oral contraceptive pill (e.g. Microgynon, Marvelon, Yasmin)
- Progestin only pill (e.g. Minipill)
- Contraceptive Injection (e.g. Depo-Provera, Sayana Press, Noristerat)
- Hormonal IUD/IUS/Coil (e.g. Mirena, Kyleena, Jaydess, Levosert or Skyla)
- Contraceptive patch (e.g. Ortho Evra, Xulane
- Vaginal ring (e.g. Nuvaring)
- Contraceptive Implant (e.g. Nexplanon)
- Copper IUD
- None of the above

Show/hide trigger exists. Hidden unless: #41 Question "
**Did you use the contraceptive patch for the entire pandemic (since 2020)?**

" is one of the following answers ("No")

56. **Did you use any hormonal method or copper IUD after you stopped using the contraceptive patch?** *Select which method you used next. If you started the same method again, select that method.*

- Combined oral contraceptive pill (e.g. Microgynon, Marvelon, Yasmin)
- Progestin only pill (e.g. Minipill)
- Contraceptive Injection (e.g. Depo-Provera, Sayana Press, Noristerat)
- Hormonal IUD/IUS/Coil (e.g. Mirena, Kyleena, Jaydess, Levosert or Skyla)
- Contraceptive patch (e.g. Ortho Evra, Xulane
- Vaginal ring (e.g. Nuvaring)
- Contraceptive Implant (e.g. Nexplanon)
- Copper IUD
- None of the above

Show/hide trigger exists. Hidden unless: #42 Question "
**Did you use the vaginal ring for the entire pandemic (since 2020)?**

" is one of the following answers ("No")

57. **Did you use any hormonal method or copper IUD after you stopped using the vaginal ring?** *Select which method you used next. If you started the same method again, select that method.*

- Combined oral contraceptive pill (e.g. Microgynon, Marvelon, Yasmin)
- Progestin only pill (e.g. Minipill)
- Contraceptive Injection (e.g. Depo-Provera, Sayana Press, Noristerat)
- Hormonal IUD/IUS/Coil (e.g. Mirena, Kyleena, Jaydess, Levosert or Skyla)
- Contraceptive patch (e.g. Ortho Evra, Xulane
- Vaginal ring (e.g. Nuvaring)
- Contraceptive Implant (e.g. Nexplanon)
- Copper IUD
- None of the above

LOGIC Show/hide trigger exists. Hidden unless: #43 Question "
**Did you use the contraceptive implant for the entire pandemic (since 2020)?**

" is one of the following answers ("No")

58. **Did you use any hormonal method or copper IUD after you stopped using the contraceptive implant?** *Select which method you used next. If you started the same method again, select that method.*

- Combined oral contraceptive pill (e.g. Microgynon, Marvelon, Yasmin)
- Progestin only pill (e.g. Minipill)
- Contraceptive Injection (e.g. Depo-Provera, Sayana Press, Noristerat)
- Hormonal IUD/IUS/Coil (e.g. Mirena, Kyleena, Jaydess, Levosert or Skyla)
- Contraceptive patch (e.g. Ortho Evra, Xulane
- Vaginal ring (e.g. Nuvaring)
- Contraceptive Implant (e.g. Nexplanon)
- Copper IUD
- None of the above

LOGIC Show/hide trigger exists. Hidden unless: #44 Question "
**Did you use the copper IUD for the entire pandemic (since 2020)?**

" is one of the following answers ("No")

59. **Did you use any hormonal method or copper IUD after you stopped using the copper IUD?** *Select which method you used next. If you started the same method again, select that method.*

- Combined oral contraceptive pill (e.g. Microgynon, Marvelon, Yasmin)
- Progestin only pill (e.g. Minipill)
- Contraceptive Injection (e.g. Depo-Provera, Sayana Press, Noristerat)
- Hormonal IUD/IUS/Coil (e.g. Mirena, Kyleena, Jaydess, Levosert or Skyla)
- Contraceptive patch (e.g. Ortho Evra, Xulane
- Vaginal ring (e.g. Nuvaring)
- Contraceptive Implant (e.g. Nexplanon)
- Copper IUD
- None of the above

LOGIC Hidden unless: #52 Question "
**Did you use any hormonal method or copper IUD after you stopped using combined oral contraceptives? Select which method you used next.** *If you started the same method again, select that method.*

" is one of the following answers ("Combined oral contraceptive pill (e.g. Microgynon, Marvelon, Yasmin)","Progestin only pill (e.g. Minipill)","Contraceptive Injection (e.g. Depo-Provera, Sayana Press, Noristerat)","Hormonal IUD/IUS/Coil (e.g. Mirena, Kyleena, Jaydess, Levosert or Skyla)","Contraceptive patch (e.g. Ortho Evra, Xulane","Vaginal ring (e.g. Nuvaring)","Contraceptive Implant (e.g. Nexplanon)","Copper IUD")

60. **When did you start using this method?**

Feb 2020
March 2020
April 2020
May 2020
June 2020
July 2020
August 2020
Sept 2020
Oct 2020
Nov 2020
Dec 2020
Jan 2021
Feb 2021
March 2021
April 2021
May 2021
June 2021
July 2021
August 2021
Sept 2021
Oct 2021
Nov 2021
Dec 2021

LOGIC Hidden unless: #52 Question "
**Did you use any hormonal method or copper IUD after you stopped using combined oral contraceptives? Select which method you used next.** *If you started the same method again, select that method.*

" is one of the following answers ("Combined oral contraceptive pill (e.g. Microgynon, Marvelon, Yasmin)","Progestin only pill (e.g. Minipill)","Contraceptive Injection (e.g. Depo-Provera, Sayana Press, Noristerat)","Hormonal IUD/IUS/Coil (e.g. Mirena, Kyleena, Jaydess, Levosert or Skyla)","Contraceptive patch (e.g. Ortho Evra, Xulane","Vaginal ring (e.g. Nuvaring)","Contraceptive Implant (e.g. Nexplanon)","Copper IUD")

61. **Did you use this method for the rest of the pandemic (until now)?**

- Yes
- No

LOGIC Hidden unless: #53 Question "
**Did you use any hormonal method or copper IUD after you stopped using the progestin-only pill?** *Select which method you used next. If you started the same method again, select that method.*

" is one of the following answers ("Combined oral contraceptive pill (e.g. Microgynon, Marvelon, Yasmin)","Progestin only pill (e.g. Minipill)","Contraceptive Injection (e.g. Depo-Provera, Sayana Press, Noristerat)","Hormonal IUD/IUS/Coil (e.g. Mirena, Kyleena, Jaydess, Levosert or Skyla)","Contraceptive patch (e.g. Ortho Evra, Xulane","Vaginal ring (e.g. Nuvaring)","Contraceptive Implant (e.g. Nexplanon)","Copper IUD")

62. **When did you start using this method?**

Feb 2020
March 2020
April 2020
May 2020
June 2020
July 2020
August 2020
Sept 2020
Oct 2020
Nov 2020
Dec 2020
Jan 2021
Feb 2021
March 2021
April 2021
May 2021
June 2021
July 2021
August 2021
Sept 2021
Oct 2021
Nov 2021
Dec 2021

LOGIC Hidden unless: #53 Question "
**Did you use any hormonal method or copper IUD after you stopped using the progestin-only pill?** *Select which method you used next. If you started the same method again, select that method.*

" is one of the following answers ("Combined oral contraceptive pill (e.g. Microgynon, Marvelon, Yasmin)","Progestin only pill (e.g. Minipill)","Contraceptive Injection (e.g. Depo-Provera, Sayana Press, Noristerat)","Hormonal IUD/IUS/Coil (e.g. Mirena, Kyleena, Jaydess, Levosert or Skyla)","Contraceptive patch (e.g. Ortho Evra, Xulane","Vaginal ring (e.g. Nuvaring)","Contraceptive Implant (e.g. Nexplanon)","Copper IUD")

63. **Did you use this method for the rest of the pandemic (until now)?**

- ○ Yes
- ○ No

LOGIC Hidden unless: #54 Question "

**Did you use any hormonal method or copper IUD after you stopped using the contraceptive injection?** *Select which method you used next. If you started the same method again, select that method.*

" is one of the following answers ("Combined oral contraceptive pill (e.g. Microgynon, Marvelon, Yasmin)","Progestin only pill (e.g. Minipill)","Contraceptive Injection (e.g. Depo-Provera, Sayana Press, Noristerat)","Hormonal IUD/IUS/Coil (e.g. Mirena, Kyleena, Jaydess, Levosert or Skyla)","Contraceptive patch (e.g. Ortho Evra, Xulane","Vaginal ring (e.g. Nuvaring)","Contraceptive Implant (e.g. Nexplanon)","Copper IUD")

64. **When did you start using this method?**

Feb 2020
March 2020
April 2020
May 2020
June 2020
July 2020
August 2020
Sept 2020
Oct 2020
Nov 2020
Dec 2020
Jan 2021
Feb 2021
March 2021
April 2021
May 2021
June 2021
July 2021
August 2021
Sept 2021
Oct 2021
Nov 2021
Dec 2021

LOGIC Hidden unless: #55 Question "

**Did you use any hormonal method or copper IUD after you stopped using the hormonal IUD/IUS/Coil?** *Select which method you used next. If you started the same method again, select that method.*

" is one of the following answers ("Combined oral contraceptive pill (e.g. Microgynon, Marvelon, Yasmin)","Progestin only pill (e.g. Minipill)","Contraceptive Injection (e.g. Depo-Provera, Sayana Press, Noristerat)","Hormonal IUD/IUS/Coil (e.g. Mirena, Kyleena, Jaydess, Levosert or Skyla)","Contraceptive patch (e.g. Ortho Evra, Xulane","Vaginal ring (e.g. Nuvaring)","Contraceptive Implant (e.g. Nexplanon)","Copper IUD")

65. **Did you use this method for the rest of the pandemic (until now)?**

- Yes
- No

LOGIC Hidden unless: #55 Question "

**Did you use any hormonal method or copper IUD after you stopped using the hormonal IUD/IUS/Coil?** *Select which method you used next. If you started the same method again, select that method.*

" is one of the following answers ("Combined oral contraceptive pill (e.g. Microgynon, Marvelon, Yasmin)","Progestin only pill (e.g. Minipill)","Contraceptive Injection (e.g. Depo-Provera, Sayana Press, Noristerat)","Hormonal IUD/IUS/Coil (e.g. Mirena, Kyleena, Jaydess, Levosert or Skyla)","Contraceptive patch (e.g. Ortho Evra, Xulane","Vaginal ring (e.g. Nuvaring)","Contraceptive Implant (e.g. Nexplanon)","Copper IUD")

66. **When did you start using this method?**

Feb 2020
March 2020
April 2020
May 2020
June 2020
July 2020
August 2020
Sept 2020
Oct 2020
Nov 2020
Dec 2020
Jan 2021
Feb 2021
March 2021
April 2021
May 2021
June 2021
July 2021
August 2021
Sept 2021
Oct 2021
Nov 2021
Dec 2021

LOGIC Hidden unless: #55 Question "

**Did you use any hormonal method or copper IUD after you stopped using the hormonal IUD/IUS/Coil?** *Select which method you used next. If you started the same method again, select that method.*

" is one of the following answers ("Combined oral contraceptive pill (e.g. Microgynon, Marvelon, Yasmin)","Progestin only pill (e.g. Minipill)","Contraceptive Injection (e.g. Depo-Provera, Sayana Press, Noristerat)","Hormonal IUD/IUS/Coil (e.g. Mirena, Kyleena, Jaydess, Levosert or Skyla)","Contraceptive patch (e.g. Ortho Evra, Xulane","Vaginal ring (e.g. Nuvaring)","Contraceptive Implant (e.g. Nexplanon)","Copper IUD")

67. **Did you use this method for the rest of the pandemic (until now)?**

- Yes
- No

LOGIC Hidden unless: #56 Question "
**Did you use any hormonal method or copper IUD after you stopped using the contraceptive patch?** *Select which method you used next. If you started the same method again, select that method.*

" is one of the following answers ("Combined oral contraceptive pill (e.g. Microgynon, Marvelon, Yasmin)","Progestin only pill (e.g. Minipill)","Contraceptive Injection (e.g. Depo-Provera, Sayana Press, Noristerat)","Hormonal IUD/IUS/Coil (e.g. Mirena, Kyleena, Jaydess, Levosert or Skyla)","Contraceptive patch (e.g. Ortho Evra, Xulane","Vaginal ring (e.g. Nuvaring)","Contraceptive Implant (e.g. Nexplanon)","Copper IUD")

68. **When did you start using this method?**

Feb 2020
March 2020
April 2020
May 2020
June 2020
July 2020
August 2020
Sept 2020
Oct 2020
Nov 2020
Dec 2020
Jan 2021
Feb 2021
March 2021
April 2021
May 2021
June 2021
July 2021
August 2021
Sept 2021
Oct 2021
Nov 2021
Dec 2021

LOGIC Hidden unless: #56 Question "
**Did you use any hormonal method or copper IUD after you stopped using the contraceptive patch?** *Select which method you used next. If you started the same method again, select that method.*

" is one of the following answers ("Combined oral contraceptive pill (e.g. Microgynon, Marvelon, Yasmin)","Progestin only pill (e.g. Minipill)","Contraceptive Injection (e.g. Depo-Provera, Sayana Press, Noristerat)","Hormonal IUD/IUS/Coil (e.g. Mirena, Kyleena, Jaydess, Levosert or Skyla)","Contraceptive patch (e.g. Ortho Evra, Xulane","Vaginal ring (e.g. Nuvaring)","Contraceptive Implant (e.g. Nexplanon)","Copper IUD")

69. **Did you use this method for the rest of the pandemic (until now)?**

- ○ Yes
- ○ No

LOGIC Hidden unless: #57 Question "

**Did you use any hormonal method or copper IUD after you stopped using the vaginal ring?** *Select which method you used next. If you started the same method again, select that method.*

" is one of the following answers ("Combined oral contraceptive pill (e.g. Microgynon, Marvelon, Yasmin)","Progestin only pill (e.g. Minipill)","Contraceptive Injection (e.g. Depo-Provera, Sayana Press, Noristerat)","Hormonal IUD/IUS/Coil (e.g. Mirena, Kyleena, Jaydess, Levosert or Skyla)","Contraceptive patch (e.g. Ortho Evra, Xulane","Vaginal ring (e.g. Nuvaring)","Contraceptive Implant (e.g. Nexplanon)","Copper IUD")

70. **When did you start using this method?**

Feb 2020
March 2020
April 2020
May 2020
June 2020
July 2020
August 2020
Sept 2020
Oct 2020
Nov 2020
Dec 2020
Jan 2021
Feb 2021
March 2021
April 2021
May 2021
June 2021
July 2021
August 2021
Sept 2021
Oct 2021
Nov 2021
Dec 2021

LOGIC Hidden unless: #57 Question "

**Did you use any hormonal method or copper IUD after you stopped using the vaginal ring?** *Select which method you used next. If you started the same method again, select that method.*

" is one of the following answers ("Combined oral contraceptive pill (e.g. Microgynon, Marvelon, Yasmin)","Progestin only pill (e.g. Minipill)","Contraceptive Injection (e.g. Depo-Provera, Sayana Press, Noristerat)","Hormonal IUD/IUS/Coil (e.g. Mirena, Kyleena, Jaydess, Levosert or Skyla)","Contraceptive patch (e.g. Ortho Evra, Xulane","Vaginal ring (e.g. Nuvaring)","Contraceptive Implant (e.g. Nexplanon)","Copper IUD")

71. **Did you use this method for the rest of the pandemic (until now)?**

- Yes
- No

LOGIC Hidden unless: #58 Question "

**Did you use any hormonal method or copper IUD after you stopped using the contraceptive implant?** *Select which method you used next. If you started the same method again, select that method.*

" is one of the following answers ("Combined oral contraceptive pill (e.g. Microgynon, Marvelon, Yasmin)","Progestin only pill (e.g. Minipill)","Contraceptive Injection (e.g. Depo-Provera, Sayana Press, Noristerat)","Hormonal IUD/IUS/Coil (e.g. Mirena, Kyleena, Jaydess, Levosert or Skyla)","Contraceptive patch (e.g. Ortho Evra, Xulane","Vaginal ring (e.g. Nuvaring)","Contraceptive Implant (e.g. Nexplanon)","Copper IUD")

72. **When did you start using this method?**

- Feb 2020
- March 2020
- April 2020
- May 2020
- June 2020
- July 2020
- August 2020
- Sept 2020
- Oct 2020
- Nov 2020
- Dec 2020
- Jan 2021
- Feb 2021
- March 2021
- April 2021
- May 2021
- June 2021
- July 2021
- August 2021
- Sept 2021
- Oct 2021
- Nov 2021
- Dec 2021

LOGIC Hidden unless: #58 Question "

**Did you use any hormonal method or copper IUD after you stopped using the contraceptive implant?** *Select which method you used next. If you started the same method again, select that method.*

" is one of the following answers ("Combined oral contraceptive pill (e.g. Microgynon, Marvelon, Yasmin)","Progestin only pill (e.g. Minipill)","Contraceptive Injection (e.g. Depo-Provera, Sayana Press, Noristerat)","Hormonal IUD/IUS/Coil (e.g. Mirena, Kyleena, Jaydess, Levosert or Skyla)","Contraceptive patch (e.g. Ortho Evra, Xulane","Vaginal ring (e.g. Nuvaring)","Contraceptive Implant (e.g. Nexplanon)","Copper IUD")

73. **Did you use this method for the rest of the pandemic (until now)?**

- ○ Yes
- ○ No

LOGIC Hidden unless: #59 Question "

**Did you use any hormonal method or copper IUD after you stopped using the copper IUD?** *Select which method you used next. If you started the same method again, select that method.*

" is one of the following answers ("Combined oral contraceptive pill (e.g. Microgynon, Marvelon, Yasmin)","Progestin only pill (e.g. Minipill)","Contraceptive Injection (e.g. Depo-Provera, Sayana Press, Noristerat)","Hormonal IUD/IUS/Coil (e.g. Mirena, Kyleena, Jaydess, Levosert or Skyla)","Contraceptive patch (e.g. Ortho Evra, Xulane","Vaginal ring (e.g. Nuvaring)","Contraceptive Implant (e.g. Nexplanon)","Copper IUD")

74. **When did you start using this method?**

- Feb 2020
- March 2020
- April 2020
- May 2020
- June 2020
- July 2020
- August 2020
- Sept 2020
- Oct 2020
- Nov 2020
- Dec 2020
- Jan 2021
- Feb 2021
- March 2021
- April 2021
- May 2021
- June 2021
- July 2021
- August 2021
- Sept 2021
- Oct 2021
- Nov 2021
- Dec 2021

LOGIC Hidden unless: #59 Question "

**Did you use any hormonal method or copper IUD after you stopped using the copper IUD?** *Select which method you used next. If you started the same method again, select that method.*

" is one of the following answers ("Combined oral contraceptive pill (e.g. Microgynon, Marvelon, Yasmin)","Progestin only pill (e.g. Minipill)","Contraceptive Injection (e.g. Depo-Provera, Sayana Press, Noristerat)","Hormonal IUD/IUS/Coil (e.g. Mirena, Kyleena, Jaydess, Levosert or Skyla)","Contraceptive patch (e.g. Ortho Evra, Xulane","Vaginal ring (e.g. Nuvaring)","Contraceptive Implant (e.g. Nexplanon)","Copper IUD")

75. **Did you use this method for the rest of the pandemic (until now)?**

- ○ Yes
- ○ No

LOGIC Show/hide trigger exists. Hidden unless: #35 Question "**Were you using a hormonal contraceptive or a copper IUDin January 2020?** " is one of the following answers ("No")

76. **Have you used any hormonal contraceptives or a copper IUD since January 2020?** Select the first method you used after January 2020.

- Combined oral contraceptive pill (e.g. Microgynon, Marvelon, Yasmin)
- Progestin only pill (e.g. Minipill)
- Contraceptive Injection (e.g. Depo-Provera, Sayana Press, Noristerat)
- Hormonal IUD/IUS/Coil (e.g. Mirena, Kyleena, Jaydess, Levosert or Skyla)
- Contraceptive patch (e.g. Ortho Evra, Xulane
- Vaginal ring (e.g. Nuvaring)
- Contraceptive Implant (e.g. Nexplanon)
- Copper IUD
- I have not used a hormonal contraceptive or copper IUD since January 2020

LOGIC Hidden unless: #76 Question "

**Have you used any hormonal contraceptives or a copper IUD since January 2020?** Select the first method you used after January 2020.

" is one of the following answers ("Combined oral contraceptive pill (e.g. Microgynon, Marvelon, Yasmin)")

77. **When did you start using the combined oral contraceptive pill?**

Jan 2020
Feb 2020
March 2020
April 2020
May 2020
June 2020
July 2020
August 2020
Sept 2020
Oct 2020
Nov 2020
Dec 2020
Jan 2021
Feb 2021
March 2021
April 2021
May 2021
June 2021
July 2021
August 2021
Sept 2021
Oct 2021
Nov 2021
Dec 2021

LOGIC Hidden unless: #76 Question "
**Have you used any hormonal contraceptives or a copper IUD since January 2020?** Select the first method you used after January 2020.

" is one of the following answers ("Combined oral contraceptive pill (e.g. Microgynon, Marvelon, Yasmin)")

78. **Have you used combined oral contraceptives for the rest of the pandemic (until now)?**

- Yes
- No

LOGIC Hidden unless: #76 Question "
**Have you used any hormonal contraceptives or a copper IUD since January 2020?** Select the first method you used after January 2020.

" is one of the following answers ("Progestin only pill (e.g. Minipill)")

79. **When did you start using the progestin-only pill?**

Jan 2020
Feb 2020
March 2020
April 2020
May 2020
June 2020
July 2020
August 2020
Sept 2020
Oct 2020
Nov 2020
Dec 2020
Jan 2021
Feb 2021
March 2021
April 2021
May 2021
June 2021
July 2021
August 2021
Sept 2021
Oct 2021
Nov 2021
Dec 2021

LOGIC Hidden unless: #76 Question "
**Have you used any hormonal contraceptives or a copper IUD since January 2020?** Select the first method you used after January 2020.

" is one of the following answers ("Progestin only pill (e.g. Minipill)")

80. **Have you used the progestin-only pill for the rest of the pandemic (until now)?**

- Yes
- No

LOGIC Hidden unless: #76 Question "
**Have you used any hormonal contraceptives or a copper IUD since January 2020?** Select the first method you used after January 2020.

" is one of the following answers ("Contraceptive Injection (e.g. Depo-Provera, Sayana Press, Noristerat)")

81. **When did you start using the contraceptive injection?**

- Jan 2020
- Feb 2020
- March 2020
- April 2020
- May 2020
- June 2020
- July 2020
- August 2020
- Sept 2020
- Oct 2020
- Nov 2020
- Dec 2020
- Jan 2021
- Feb 2021
- March 2021
- April 2021
- May 2021
- June 2021
- July 2021
- August 2021
- Sept 2021
- Oct 2021
- Nov 2021
- Dec 2021

LOGIC Hidden unless: #76 Question "
**Have you used any hormonal contraceptives or a copper IUD since January 2020?** Select the first method you used after January 2020.

" is one of the following answers ("Contraceptive Injection (e.g. Depo-Provera, Sayana Press, Noristerat)")

82. **Have you used the contraceptive injection for the rest of the pandemic (until now)?**

- Yes
- No

LOGIC Hidden unless: #76 Question "
**Have you used any hormonal contraceptives or a copper IUD since January 2020?** Select the first method you used after January 2020.

" is one of the following answers ("Hormonal IUD/IUS/Coil (e.g. Mirena, Kyleena, Jaydess, Levosert or Skyla)")

83. **When did you start using the hormonal IUS/IUD/Coil?**

- Jan 2020
- Feb 2020
- March 2020
- April 2020
- May 2020
- June 2020
- July 2020
- August 2020
- Sept 2020
- Oct 2020
- Nov 2020
- Dec 2020
- Jan 2021
- Feb 2021
- March 2021
- April 2021
- May 2021
- June 2021
- July 2021
- August 2021
- Sept 2021
- Oct 2021
- Nov 2021
- Dec 2021

LOGIC Hidden unless: #76 Question "
**Have you used any hormonal contraceptives or a copper IUD since January 2020?** Select the first method you used after January 2020.

" is one of the following answers ("Hormonal IUD/IUS/Coil (e.g. Mirena, Kyleena, Jaydess, Levosert or Skyla)")

84. **Have you used the hormonal IUD/IUS/Coil for the rest of the pandemic (until now)?**

- ○ Yes
- ○ No

LOGIC Hidden unless: #76 Question "
**Have you used any hormonal contraceptives or a copper IUD since January 2020?** Select the first method you used after January 2020.

" is one of the following answers ("Contraceptive patch (e.g. Ortho Evra, Xulane")

85. **When did you start using the contraceptive patch?**

- Jan 2020
- Feb 2020
- March 2020
- April 2020
- May 2020
- June 2020
- July 2020
- August 2020
- Sept 2020
- Oct 2020
- Nov 2020
- Dec 2020
- Jan 2021
- Feb 2021
- March 2021
- April 2021
- May 2021
- June 2021
- July 2021
- August 2021
- Sept 2021
- Oct 2021
- Nov 2021
- Dec 2021

LOGIC Hidden unless: #76 Question "
**Have you used any hormonal contraceptives or a copper IUD since January 2020?** Select the first method you used after January 2020.

" is one of the following answers ("Contraceptive patch (e.g. Ortho Evra, Xulane")

86. **Have you used the contraceptive patch for the rest of the pandemic (until now)?**

- Yes
- No

LOGIC Hidden unless: #76 Question "
**Have you used any hormonal contraceptives or a copper IUD since January 2020?** Select the first method you used after January 2020.

" is one of the following answers ("Vaginal ring (e.g. Nuvaring)")

87. **When did you start using the vaginal ring?**

- Jan 2020
- Feb 2020
- March 2020
- April 2020
- May 2020
- June 2020
- July 2020
- August 2020
- Sept 2020
- Oct 2020
- Nov 2020
- Dec 2020
- Jan 2021
- Feb 2021
- March 2021
- April 2021
- May 2021
- June 2021
- July 2021
- August 2021
- Sept 2021
- Oct 2021
- Nov 2021
- Dec 2021

LOGIC Hidden unless: #76 Question "
**Have you used any hormonal contraceptives or a copper IUD since January 2020?** Select the first method you used after January 2020.

" is one of the following answers ("Vaginal ring (e.g. Nuvaring)")

88. **Have you used the vaginal ring for the rest of the pandemic (until now)?**

- Yes
- No

LOGIC Hidden unless: #76 Question "
**Have you used any hormonal contraceptives or a copper IUD since January 2020?** Select the first method you used after January 2020.

" is one of the following answers ("Contraceptive Implant (e.g. Nexplanon)")

89. **When did you start using the contraceptive implant?**

- Jan 2020
- Feb 2020
- March 2020
- April 2020
- May 2020
- June 2020
- July 2020
- August 2020
- Sept 2020
- Oct 2020
- Nov 2020
- Dec 2020
- Jan 2021
- Feb 2021
- March 2021
- April 2021
- May 2021
- June 2021
- July 2021
- August 2021
- Sept 2021
- Oct 2021
- Nov 2021
- Dec 2021

LOGIC Hidden unless: #76 Question "
**Have you used any hormonal contraceptives or a copper IUD since January 2020?** Select the first method you used after January 2020.

" is one of the following answers ("Contraceptive Implant (e.g. Nexplanon)")

90. **Have you used the contraceptive implant for the rest of the pandemic (until now)?**

- Yes
- No

Hidden unless: #76 Question "
**Have you used any hormonal contraceptives or a copper IUD since January 2020?** Select the first method you used after January 2020.

" is one of the following answers ("Copper IUD")

91. **When did you start using the copper IUD?**

- Jan 2020
- Feb 2020
- March 2020
- April 2020
- May 2020
- June 2020
- July 2020
- August 2020
- Sept 2020
- Oct 2020
- Nov 2020
- Dec 2020
- Jan 2021
- Feb 2021
- March 2021
- April 2021
- May 2021
- June 2021
- July 2021
- August 2021
- Sept 2021
- Oct 2021
- Nov 2021
- Dec 2021

Hidden unless: #76 Question "
**Have you used any hormonal contraceptives or a copper IUD since January 2020?** Select the first method you used after January 2020.

" is one of the following answers ("Copper IUD")

92. **Have you used the copper IUD for the rest of the pandemic (until now)?**

- Yes
- No

Show/hide trigger exists.

93. **Did you undergo any gynecologic surgeries/procedures since the beginning of 2020?**

- Yes
- No

LOGIC Hidden unless: #93 Question "**Did you undergo any gynecologic surgeries/procedures since the beginning of 2020?** is one of the following answers ("Yes")

94. **What were the surgical procedures you underwent?** (check all that apply)

- ☐ Endometrial biopsy (a small piece of the uterus is removed)
- ☐ Myomectomy (fibroid removal surgery)
- ☐ Dilation and curettage (surgical miscarriage or abortion)
- ☐ Polypectomy (removal of polyp)
- ☐ Hysteroscopy (removal of uterus)
- ☐ Loop electrosurgical excision procedure (LEEP, to remove precancerous cells from the cervix)
- ☐ None of the above

LOGIC Hidden unless: #93 Question "**Did you undergo any gynecologic surgeries/procedures since the beginning of 2020?** is one of the following answers ("Yes")

95. **When was your gynecologic procedure?** (If you have undergone several procedures, select the date of the first one)

Before 2020
Jan 2020
Feb 2020
Mar 2020
Apr 2020
May 2020
June 2020
Jul 2020
Aug 2020
Sept 2020
Oct 2020
Nov 2020
Dec 2020
Jan 2021
Feb 2021
Mar 2021
Apr 2021
May 2021
June 2021
July 2021
August 2021
September 2021
October 2021
Nov 2021
Dec 2021

96. **Has your weight changed since the beginning of 2020?**

- ○ Yes, I have gained weight
- ○ Yes, I have lost weight
- ○ Yes, I lost and gained weight at different times
- ○ No, I am about the same weight
- ○ Prefer not to say

97. **Which of the following have you experienced during the pandemic?** (check all that apply)

- ☐ Loss of loved ones (e.g. bereavement)
- ☐ A close family member or friend had to be hospitalized because of serious illness or injury
- ☐ Serious illness or injury requiring my hospitalization
- ☐ Increased stigma or discrimination from other people
- ☐ Personal financial loss
- ☐ Not having enough basic supplies (e.g., food, water, medications, a place to stay)
- ☐ Anxiety / depression
- ☐ Changes to my typical sleep pattern
- ☐ Increased alcohol or other substance use
- ☐ Loneliness
- ☐ Fear
- ☐ Worry for friends and family
- ☐ None of the above

98. **In total, how much time have you spent in isolation or quarantine because of lockdowns or travel measures, and/or following possible COVID-19 exposure or positive COVID-19 test(s)?**

- ○ None
- ○ Less than a week
- ○ 1-4 weeks
- ○ 5-8 weeks
- ○ 9-12 weeks
- ○ More than 12 weeks

99. **Did you have to balance working from home and taking care of others (e.g., parents, kids, partners) during the pandemic?**

- ○ Yes
- ○ No

100. **How comfortable was your home/living space situation during the pandemic?** (in terms of space available for yourself)

- Very uncomfortable
- Slightly uncomfortable
- Neither comfortable / uncomfortable
- Slightly comfortable
- Very comfortable

101. When you were most restricted due to the pandemic, did you have access to any of the following outdoor spaces from your home?

- [ ] Private or shared garden / yard
- [ ] Balcony / terrace
- [ ] Other accessible spaces (e.g. park nearby)
- [ ] None of the above

102. **On a scale from 0-10, how stressed have you been feeling, on average, since the beginning of the pandemic?**

Not stressed ——— Extremely stressed

---

LOGIC Show/hide trigger exists.

103. **Have you ever been infected with COVID-19?** *

- Yes
- No
- I'm not sure
- Prefer not to answer

Show/hide trigger exists. Hidden unless: #103 Question "
**Have you ever been infected with COVID-19?**

" is one of the following answers ("Yes")

104. **Did you experience any symptoms with COVID-19?**

- I did not experience symptoms
- I had some mild cold symptoms but they didn’t interfere with my activities
- I had moderate symptoms that interfered with my activities
- I had moderate symptoms that prevented me from my activities
- I had severe symptoms and I had to get emergency care but I did not need to stay in the hospital
- I had severe symptoms and I needed oxygen therapy in the hospital but I did not need a ventilator
- I had severe symptoms and I needed a ventilator

Hidden unless: #104 Question "**Did you experience any symptoms with COVID-19?**" is one of the following answers ("I had some mild cold symptoms but they didn’t interfere with my activities","I had moderate symptoms that interfered with my activities","I had moderate symptoms that prevented me from my activities","I had severe symptoms and I had to get emergency care but I did not need to stay in the hospital","I had severe symptoms and I needed oxygen therapy in the hospital but I did not need a ventilator","I had severe symptoms and I needed a ventilator")

105. **When did your first symptoms start?**

I don't know
January 2020
February 2020
March 2020
April 2020
May 2020
June 2020
July 2020
August 2020
September 2020
October 2020
November 2020
December 2020
January 2021
February 2021
March 2021
April 2021
May 2021
June 2021
July 2021
August 2021
September 2021
October 2021
November 2021
December 2021

LOGIC Show/hide trigger exists. Hidden unless: #103 Question "
**Have you ever been infected with COVID-19?**

" is one of the following answers ("Yes")

106. **Have you had at least one positive test for COVID-19?** *

- Yes
- No but I have had a clinical diagnosis by a doctor based on symptoms
- No

LOGIC Hidden unless: #106 Question "
**Have you had at least one positive test for COVID-19?**

" is one of the following answers ("Yes")

107. **When was your first positive test?**

- I don't know
- January 2020
- February 2020
- March 2020
- April 2020
- May 2020
- June 2020
- July 2020
- August 2020
- September 2020
- October 2020
- November 2020
- December 2020
- January 2021
- February 2021
- March 2021
- April 2021
- May 2021
- June 2021
- July 2021
- August 2021
- September 2021
- October 2021
- November 2021
- December 2021

LOGIC Hidden unless: #106 Question "
**Have you had at least one positive test for COVID-19?**

" is one of the following answers ("No but I have had a clinical diagnosis by a doctor based on symptoms")

108. **When were you diagnosed?**

- I don't know
- January 2020
- February 2020
- March 2020
- April 2020
- May 2020
- June 2020
- July 2020
- August 2020
- September 2020
- October 2020
- November 2020
- December 2020
- January 2021
- February 2021
- March 2021
- April 2021
- May 2021
- June 2021
- July 2021
- August 2021
- September 2021
- October 2021
- November 2021
- December 2021

LOGIC Hidden unless: #106 Question "
**Have you had at least one positive test for COVID-19?**

" is one of the following answers ("Yes")

109. **What was the type of the test that came back positive?**

- PCR test (nose/throat swab)
- Antigen / Rapid test
- I don't know which type of test but it was a nose/throat swab
- Antibody blood test
- Other (please specify)

LOGIC Show/hide trigger exists. Hidden unless: #103 Question "
**Have you ever been infected with COVID-19?**

" is one of the following answers ("Yes")

110. **Have you experienced long-haul COVID-19 (symptoms lasting more 1 month)?** *

- Yes
- No
- I'm not sure
- Prefer not to answer

LOGIC Hidden unless: #110 Question "
**Have you experienced long-haul COVID-19 (symptoms lasting more 1 month)?**

" is one of the following answers ("Yes")

111. **Are you still experiencing symptoms from your COVID-19 infection?**

- Yes
- No

LOGIC Hidden unless: #110 Question "
**Have you experienced long-haul COVID-19 (symptoms lasting more 1 month)?**

" is one of the following answers ("Yes")

112. **How would you rate how you feel today, on a scale of 0-100% (with 100% being how you felt before COVID-19)?**

1 ______________________________ 100

---

LOGIC Hidden unless: #103 Question "
**Have you ever been infected with COVID-19?**

" is one of the following answers ("Yes")

113. **Which of the following symptoms did you experience from your COVID-19 infection (list 1)?** (check all that apply)

- Brain fog / cognitive dysfunction
- Memory impairment
- Speech and language symptoms (e.g. difficulty finding words)
- Sensorimotor symptoms (e.g. tingling/pins and needles)
- Dizziness and balance issues
- Change in smell and taste
- None of the above

Hidden unless: #103 Question "
**Have you ever been infected with COVID-19?**

" is one of the following answers ("Yes")

114. **Which of the following symptoms did you experience from your COVID-19 infection (list 2)?** (check all that apply)

- ☐ Insomnia (unable to sleep)
- ☐ Headache
- ☐ Disturbed sleep
- ☐ Fatigue
- ☐ Exhaustion after exercise/effort
- ☐ Chills, flushing, sweats
- ☐ None of the above

Hidden unless: #103 Question "
**Have you ever been infected with COVID-19?**

" is one of the following answers ("Yes")

115. **Which of the following symptoms did you experience from your COVID-19 infection (list 3)?** (check all that apply)

- ☐ Fever (>98.6F, >37C)
- ☐ Heart palpitations (extra awareness of heart beat or irregular heart beat)
- ☐ Tachycardia (heart beating too fast)
- ☐ Pain/burning in the chest
- ☐ Chest tightness
- ☐ Muscle aches
- ☐ None of the above

Hidden unless: #103 Question "
**Have you ever been infected with COVID-19?**

" is one of the following answers ("Yes")

116. **Which of the following symptoms did you experience from your COVID-19 infection (list 4)?** (check all that apply)

- ☐ Joint pain
- ☐ Sore throat
- ☐ Vision symptoms (e.g. blurred vision, flashing)
- ☐ Ringing in the ears
- ☐ Shortness of breath
- ☐ Dry cough
- ☐ None of the above

Hidden unless: #103 Question "
**Have you ever been infected with COVID-19?**

" is one of the following answers ("Yes")

117. **Which of the following symptoms did you experience from your COVID-19 infection (list 5)?** (check all that apply)

- ☐ Breathing difficulty
- ☐ Diarrhea
- ☐ Loss of appetite
- ☐ Nausea
- ☐ Abdominal pain
- ☐ None of the above

---

**Page exit logic:** Skip / Disqualify Logic
**IF:** #1 Question "
**Please confirm that you have read the information above and agree with the following statements:**

" is not one of the following answers ("I have read the information and voluntarily agree to participate in this study","I am 18 years of age or older","I experience menstrual periods (or withdrawal bleeds)","I have not been pregnant or breastfeeding since January 2020") **THEN:** Jump to page 9 - Thank You!

LOGIC Show/hide trigger exists.

118. **Have you been vaccinated against COVID-19?**

- No
- Yes, one shot
- Yes, two shots
- Yes, three shots
- I am not sure because I am currently enrolled in a COVID-19 clinical trial

LOGIC Hidden unless: #118 Question "

**Have you been vaccinated against COVID-19?**

" is one of the following answers ("Yes, one shot","Yes, two shots","Yes, three shots")

119. **Which type of vaccine did you have for your first dose?**

- Pfizer-BioNTech
- Johnson & Johnson
- Moderna
- Oxford-AstraZeneca
- Sinopharm
- I am not sure
- Other - Write In

LOGIC Show/hide trigger exists. Hidden unless: #118 Question "

**Have you been vaccinated against COVID-19?**

" is one of the following answers ("Yes, one shot","Yes, two shots","Yes, three shots")

120. **When did you get your first dose?**

- Before 2021
- January 2021
- February 2021
- March 2021
- April 2021
- May 2021
- June 2021
- July 2021
- August 2021
- September 2021
- October 2021
- November 2021
- December 2021
- I don't know

LOGIC Show/hide trigger exists. Hidden unless: #120 Question "**When did you get your first dose?**" is one of the following answers ("January 2021","February 2021","March 2021","April 2021","May 2021","June 2021","July 2021","August 2021","September 2021")

121. **What day of the month were you vaccinated?**

- I don't know
- 1
- 2
- 3
- 4
- 5
- 6
- 7
- 8
- 9
- 10
- 11
- 12
- 13
- 14
- 15
- 16
- 17
- 18
- 19
- 20
- 21
- 22
- 23
- 24
- 25
- 26
- 27
- 28
- 29
- 30
- 31

LOGIC Hidden unless: #121 Question "**What day of the month were you vaccinated?**" is one of the following answers ("1","2","3","4","5","6","7","8","9","10","11","12","13","14","15","16","17","18","19","20","21","22","23","24","25","26","27","28","29","30","31")

122. **How certain are you of your vaccination date?**

- Very certain
- Certain +/- 3 days
- Not certain

Show/hide trigger exists. Hidden unless: #118 Question "
**Have you been vaccinated against COVID-19?**

" is one of the following answers ("Yes, one shot","Yes, two shots","Yes, three shots")

123. **What was your overall experience with your first vaccine dose?**

- No side-effects
- I had some mild symptoms but they didn't interfere with my activities
- I had symptoms that interfered with my activities
- I had symptoms that prevented me from my activities
- I had to get emergency care or go to hospital for my symptoms

Show/hide trigger exists. Hidden unless: #123 Question "**What was your overall experience with your first vaccine dose?**" is one of the following answers ("I had some mild symptoms but they didn't interfere with my activities","I had symptoms that interfered with my activities","I had symptoms that prevented me from my activities","I had to get emergency care or go to hospital for my symptoms")

124. **Which symptoms did you experience from your first vaccine dose?** (check all that apply)

- Changes in periods / cycles
- Pain at the injection site
- Chills / Fever
- Dry cough
- Tiredness
- Aches and pains
- Sore throat
- Diarrhea
- Conjunctivitis (pink eye)
- Headache
- Loss of taste or smell
- A rash on skin, or discolouration of fingers or toes
- Difficulty breathing or shortness of breath
- Chest pain or pressure
- Loss of speech or movement
- Other (please specify)

LOGIC Hidden unless: #124 Question "**Which symptoms did you experience from your first vaccine dose?** (check all that apply)" is one of the following answers ("Changes in periods / cycles")

125. **Which of the following menstrual changes did you experience?** (check all that apply)

**Bleeding**

- [ ] Heavier
- [ ] Lighter
- [ ] Spotting

**Cycles (length of time between periods)**

- [ ] Shorter
- [ ] Longer
- [ ] Become irregular

**Period (bleeding days)**

- [ ] Longer
- [ ] Shorter
- [ ] Skip / Stopped

**Other**

- [ ] Other (please specify)

LOGIC Hidden unless: #118 Question "
**Have you been vaccinated against COVID-19?**

" is one of the following answers ("Yes, two shots","Yes, three shots")

126. **Which type of vaccine did you have for your second dose?**

- ( ) Pfizer-BioNTech
- ( ) Johnson & Johnson
- ( ) Moderna
- ( ) Oxford-AstraZeneca
- ( ) Sinopharm
- ( ) Other (please specify)
- ( ) I am not sure

LOGIC Show/hide trigger exists. Hidden unless: #118 Question "
**Have you been vaccinated against COVID-19?**

" is one of the following answers ("Yes, two shots","Yes, three shots")

127. **When did you get your second dose?**

- Before 2021
- January 2021
- February 2021
- March 2021
- April 2021
- May 2021
- June 2021
- July 2021
- August 2021
- September 2021
- October 2021
- November 2021
- December 2021
- I don't know

LOGIC Show/hide trigger exists. Hidden unless: #127 Question "**When did you get your second dose?**" is one of the following answers ("January 2021","February 2021","March 2021","April 2021","May 2021","June 2021","July 2021","August 2021","September 2021")

128. **What day of the month were you vaccinated?**

- I don't know
- 1
- 2
- 3
- 4
- 5
- 6
- 7
- 8
- 9
- 10
- 11
- 12
- 13
- 14
- 15
- 16
- 17
- 18
- 19
- 20
- 21
- 22
- 23
- 24
- 25
- 26
- 27
- 28
- 29
- 30
- 31

LOGIC Hidden unless: #128 Question "**What day of the month were you vaccinated?**" is one of the following answers ("1","2","3","4","5","6","7","8","9","10","11","12","13","14","15","16","17","18","19","20","21","22","23","24","25","26","27","28","29","30","31")

129. **How certain are you of your vaccination date?** *

- Very certain
- Certain +/- 3 days
- Not certain

LOGIC Show/hide trigger exists. Hidden unless: #118 Question "

**Have you been vaccinated against COVID-19?**

" is one of the following answers ("Yes, two shots","Yes, three shots")

130. **What was your overall experience with your second vaccine dose?**

- No side-effects
- I had some mild symptoms but they didn't interfere with my activities
- I had symptoms that interfered with my activities
- I had symptoms that prevented me from my activities
- I had to get emergency care or go to hospital for my symptoms

LOGIC Show/hide trigger exists. Hidden unless: #130 Question "**What was your overall experience with your second vaccine dose?** " is one of the following answers ("I had some mild symptoms but they didn’t interfere with my activities","I had symptoms that interfered with my activities","I had symptoms that prevented me from my activities","I had to get emergency care or go to hospital for my symptoms")

131. **Which symptoms did you experience from your second vaccine dose?** (check all that apply)

- ☐ Changes in periods / cycles
- ☐ Pain at the injection site
- ☐ Fever
- ☐ Dry cough
- ☐ Tiredness
- ☐ Aches and pains
- ☐ Sore throat
- ☐ Diarrhea
- ☐ Conjunctivitis
- ☐ Headache
- ☐ Loss of taste or smell
- ☐ A rash on skin, or discolouration of fingers or toes
- ☐ Difficulty breathing or shortness of breath
- ☐ Chest pain or pressure
- ☐ Loss of speech or movement
- ☐ Other (please specify)

Hidden unless: #131 Question "**Which symptoms did you experience from your second vaccine dose?** (check all that apply)" is one of the following answers ("Changes in periods / cycles")

132. **Which of the following menstrual changes did you experience?** (check all that apply)

**Bleeding**

- [ ] Heavier
- [ ] Lighter
- [ ] Spotting

**Cycles (length of time between periods)**

- [ ] Shorter
- [ ] Longer
- [ ] Become irregular

**Period (bleeding days)**

- [ ] Longer
- [ ] Shorter
- [ ] Skip / Stopped

**Other**

- [ ] Other (please specify)

Hidden unless: #118 Question "

**Have you been vaccinated against COVID-19?**

" is one of the following answers ("Yes, three shots")

133. **Which type of vaccine did you have for third vaccine dose?**

- ○ Pfizer-BioNTech
- ○ Johnson & Johnson
- ○ Moderna
- ○ Oxford-AstraZeneca
- ○ Sinopharm
- ○ Other (please specify)
- ○ I am not sure

Show/hide trigger exists. Hidden unless: #118 Question "
**Have you been vaccinated against COVID-19?**

" is one of the following answers ("Yes, three shots")

134. **When did you get your third dose?**

- Before 2021
- January 2021
- February 2021
- March 2021
- April 2021
- May 2021
- June 2021
- July 2021
- August 2021
- September 2021
- October 2021
- November 2021
- December 2021
- I don't know

Show/hide trigger exists. Hidden unless: #134 Question "**When did you get your third dose?**" is one of the following answers ("January 2021","February 2021","March 2021","April 2021","May 2021","June 2021","July 2021","August 2021","September 2021")

135. **What day of the month were you vaccinated?**

- I don't know
- 1
- 2
- 3
- 4
- 5
- 6
- 7
- 8
- 9
- 10
- 11
- 12
- 13
- 14
- 15
- 16
- 17
- 18
- 19
- 20
- 21
- 22
- 23
- 24
- 25
- 26
- 27
- 28
- 29
- 30
- 31

LOGIC Hidden unless: #135 Question "**What day of the month were you vaccinated?**" is one of the following answers ("1","2","3","4","5","6","7","8","9","10","11","12","13","14","15","16","17","18","19","20","21","22","23","24","25","26","27","28","29","30","31")

136. **How certain are you of your vaccination date?** *

- Very certain
- Certain +/- 3 days
- Not certain

LOGIC Show/hide trigger exists. Hidden unless: #118 Question "

**Have you been vaccinated against COVID-19?**

" is one of the following answers ("Yes, three shots")

137. **What was your overall experience with your third vaccine dose?**

- No side-effects
- I had some mild symptoms but they didn’t interfere with my activities
- I had symptoms that interfered with my activities
- I had symptoms that prevented me from my activities
- I had to get emergency care or go to hospital for my symptoms

LOGIC Show/hide trigger exists. Hidden unless: #137 Question "**What was your overall experience with your third vaccine dose?** " is one of the following answers ("I had some mild symptoms but they didn’t interfere with my activities","I had symptoms that interfered with my activities","I had symptoms that prevented me from my activities","I had to get emergency care or go to hospital for my symptoms")

138. **Which symptoms did you experience from your third vaccine dose?** (check all that apply)

- ☐ Changes in periods / cycles
- ☐ Pain at the injection site
- ☐ Fever
- ☐ Dry cough
- ☐ Tiredness
- ☐ Aches and pains
- ☐ Sore throat
- ☐ Diarrhea
- ☐ Conjunctivitis
- ☐ Headache
- ☐ Loss of taste or smell
- ☐ A rash on skin, or discolouration of fingers or toes
- ☐ Difficulty breathing or shortness of breath
- ☐ Chest pain or pressure
- ☐ Loss of speech or movement
- ☐ Other (please specify)

LOGIC Hidden unless: #138 Question "**Which symptoms did you experience from your third vaccine dose?** (check all that apply)" is one of the following answers ("Changes in periods / cycles")

139. **Which of the following menstrual changes did you experience?** (check all that apply)

**Bleeding**

- ☐ Heavier
- ☐ Lighter
- ☐ Spotting

**Cycles (length of time between periods)**

- ☐ Shorter
- ☐ Longer
- ☐ Become irregular

**Period (bleeding days)**

- ☐ Longer
- ☐ Shorter
- ☐ Skip / Stopped

**Other**

- ☐ Other (please specify) ____________

140. **That's it! Do you have any feedback for us?**

____________

New Hidden Value **Action: Hidden Value**
**Value:** Populates with the **length of time** since the survey taker started the survey
