## Supplementary figures and images for "Menstrual cycle phase and its association with COVID-19 vaccine side effects and subsequent infection: A study of period tracking app users"

### Supplementary Information 2

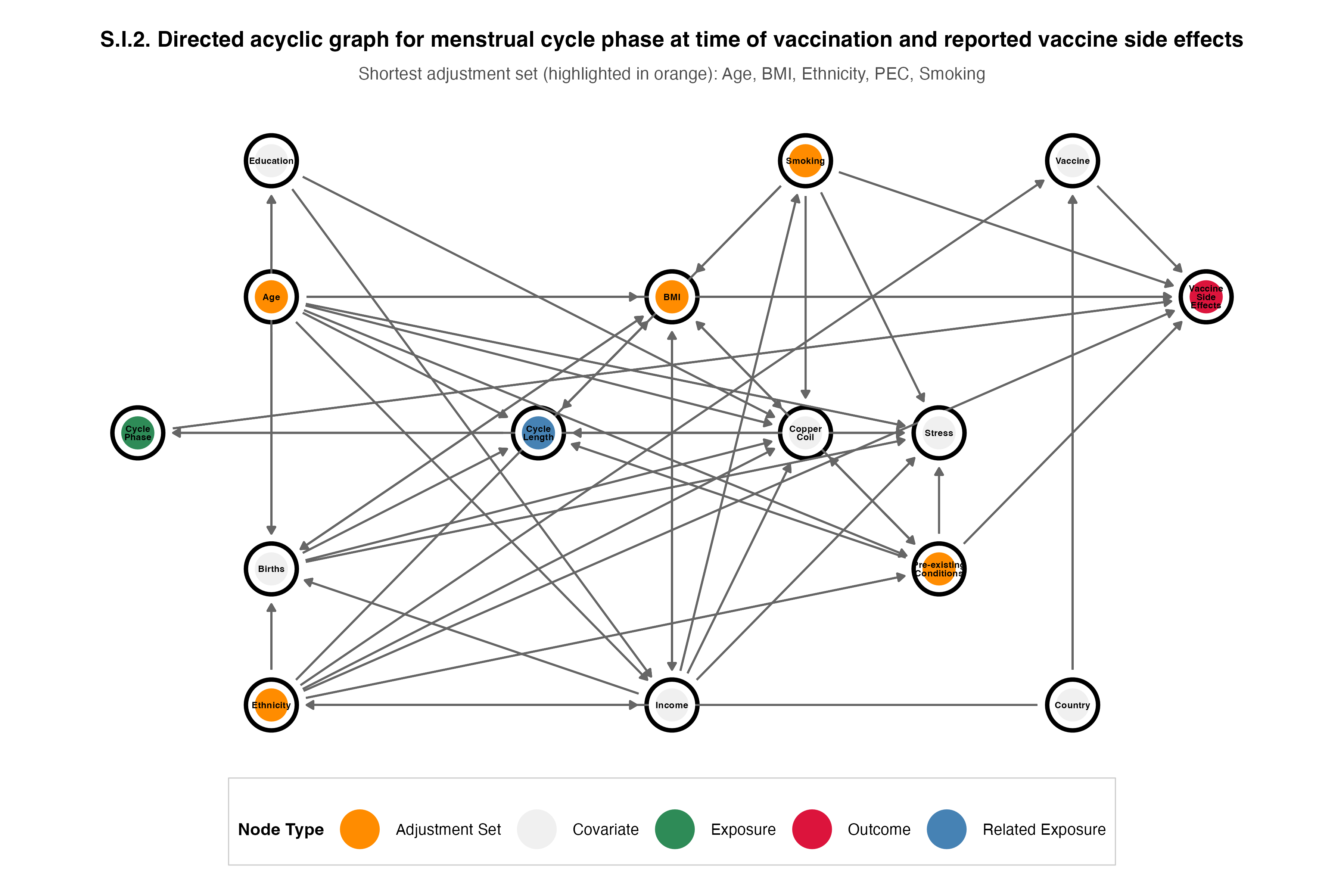

### Supplementary Information 3

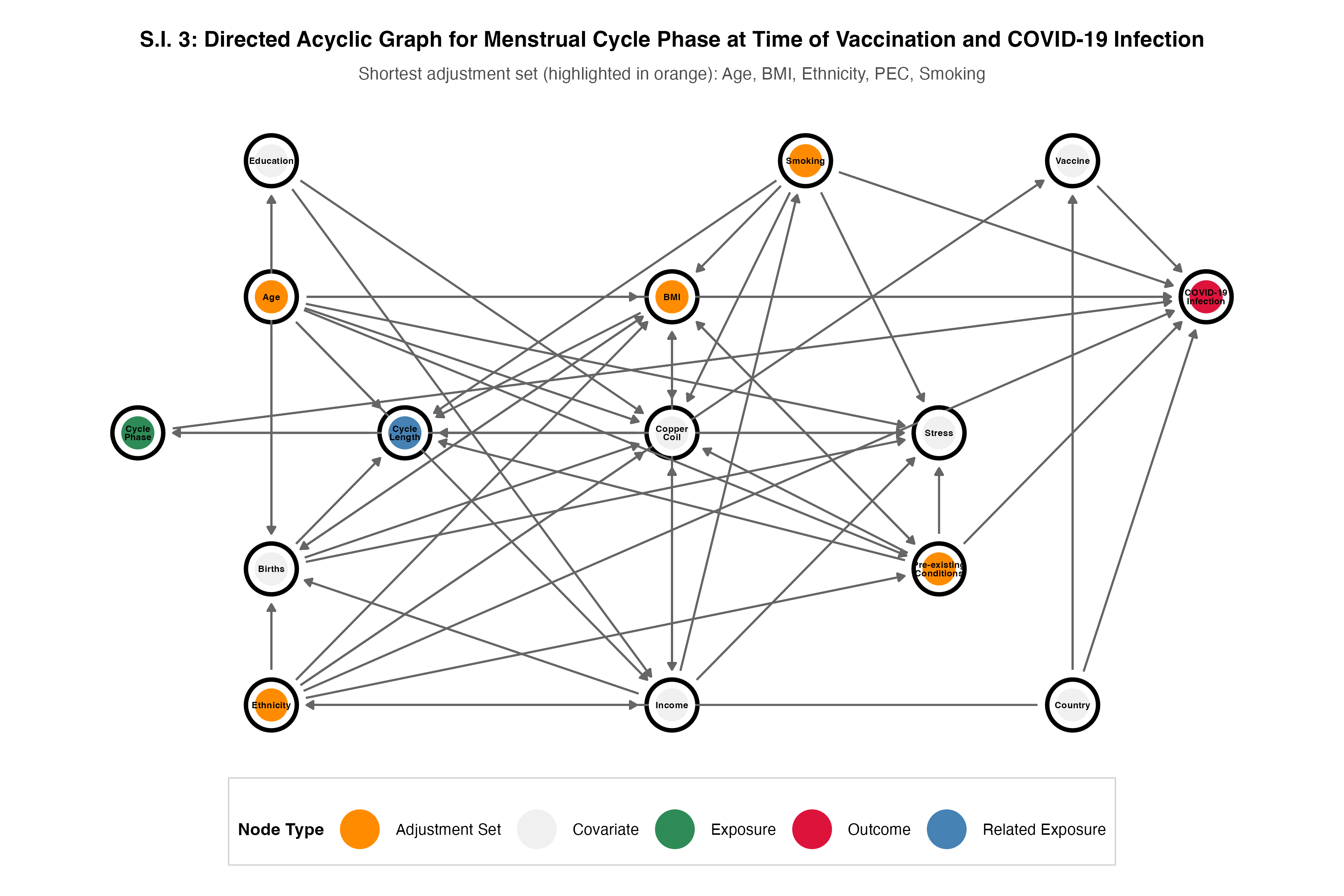
