## Supplementary Information 4 for "Menstrual cycle phase and its association with COVID-19 vaccine side effects and subsequent infection: A study of period tracking app users"

S.I.4. Participant characteristics by menstrual cycle phase at first COVID-19 vaccination: un-collapsed variables

| **Characteristic** | **N** | **Overall**  N = 1,474^1^ | **Follicular**  N = 760^1^ | **Luteal**  N = 714^1^ | **p-value**^2^ |
| --- | --- | --- | --- | --- | --- |
| **Age (years)** | 1,474 |  |  |  | 0.660 |
| 18-24 |  | 344 (23%) | 175 (23%) | 169 (24%) |  |
| 25-34 |  | 639 (43%) | 338 (44%) | 301 (42%) |  |
| 35-44 |  | 491 (33%) | 247 (33%) | 244 (34%) |  |
| **BMI category** | 1,474 |  |  |  | 0.237 |
| Healthy weight |  | 706 (48%) | 365 (48%) | 341 (48%) |  |
| Obese |  | 429 (29%) | 209 (28%) | 220 (31%) |  |
| Overweight |  | 339 (23%) | 186 (24%) | 153 (21%) |  |
| **Residence** | 1,474 |  |  |  | 0.224 |
| Australia |  | 88 (6.0%) | 39 (5.1%) | 49 (6.9%) |  |
| Canada |  | 8 (0.5%) | 2 (0.3%) | 6 (0.8%) |  |
| United Kingdom |  | 332 (23%) | 175 (23%) | 157 (22%) |  |
| United States of America |  | 1,046 (71%) | 544 (72%) | 502 (70%) |  |
| **Ethnicity** | 1,474 |  |  |  | **0.010** |
| Asian |  | 91 (6.2%) | 51 (6.7%) | 40 (5.6%) |  |
| Black |  | 62 (4.2%) | 24 (3.2%) | 38 (5.3%) |  |
| Hispanic |  | 89 (6.0%) | 42 (5.5%) | 47 (6.6%) |  |
| Middle Eastern |  | 6 (0.4%) | 5 (0.7%) | 1 (0.1%) |  |
| Mixed |  | 113 (7.7%) | 55 (7.2%) | 58 (8.1%) |  |
| Other |  | 18 (1.2%) | 6 (0.8%) | 12 (1.7%) |  |
| Prefer not to say |  | 8 (0.5%) | 8 (1.1%) | 0 (0%) |  |
| White |  | 1,087 (74%) | 569 (75%) | 518 (73%) |  |
| **Education** | 1,474 |  |  |  | 0.253 |
| High school or less |  | 105 (7.1%) | 48 (6.3%) | 57 (8.0%) |  |
| More than high school |  | 1,369 (93%) | 712 (94%) | 657 (92%) |  |
| **Income** | 1,474 |  |  |  | 0.881 |
| Don't know |  | 95 (6.4%) | 47 (6.2%) | 48 (6.7%) |  |
| Higher income |  | 465 (32%) | 243 (32%) | 222 (31%) |  |
| Lower income |  | 341 (23%) | 180 (24%) | 161 (23%) |  |
| Middle income |  | 573 (39%) | 290 (38%) | 283 (40%) |  |
| **Stress** | 1,474 |  |  |  | 0.643 |
| Average |  | 1,072 (73%) | 547 (72%) | 525 (74%) |  |
| High |  | 185 (13%) | 103 (14%) | 82 (11%) |  |
| Low |  | 157 (11%) | 78 (10%) | 79 (11%) |  |
| No Answer |  | 60 (4.1%) | 32 (4.2%) | 28 (3.9%) |  |
| **Medical conditions** | 1,474 |  |  |  | 0.813 |
| Abnormal Pap Smear |  | 112 (7.6%) | 61 (8.0%) | 51 (7.1%) |  |
| Diabetes |  | 10 (0.7%) | 4 (0.5%) | 6 (0.8%) |  |
| Eating Disorder |  | 49 (3.3%) | 22 (2.9%) | 27 (3.8%) |  |
| Endometriosis |  | 19 (1.3%) | 10 (1.3%) | 9 (1.3%) |  |
| Gynecological Cancers |  | 1 (<0.1%) | 0 (0%) | 1 (0.1%) |  |
| Immunosuppressive Therapy Condition |  | 7 (0.5%) | 3 (0.4%) | 4 (0.6%) |  |
| Interstitial Cystitis |  | 8 (0.5%) | 5 (0.7%) | 3 (0.4%) |  |
| Multiple |  | 92 (6.2%) | 50 (6.6%) | 42 (5.9%) |  |
| None |  | 1,050 (71%) | 541 (71%) | 509 (71%) |  |
| Other Cancers |  | 8 (0.5%) | 3 (0.4%) | 5 (0.7%) |  |
| PCOS |  | 40 (2.7%) | 25 (3.3%) | 15 (2.1%) |  |
| Pelvic Inflammatory Disease |  | 1 (<0.1%) | 1 (0.1%) | 0 (0%) |  |
| Thyroid |  | 56 (3.8%) | 26 (3.4%) | 30 (4.2%) |  |
| Uterine Polyps/Fibroids |  | 21 (1.4%) | 9 (1.2%) | 12 (1.7%) |  |
| **Copper IUD** | 1,474 |  |  |  | 0.821 |
| Copper IUD fitted |  | 142 (9.6%) | 75 (9.9%) | 67 (9.4%) |  |
| Not fitted |  | 1,332 (90%) | 685 (90%) | 647 (91%) |  |
| **Smoking status** | 1,474 |  |  |  | 0.160 |
| Never |  | 1,034 (70%) | 520 (68%) | 514 (72%) |  |
| Often |  | 167 (11%) | 85 (11%) | 82 (11%) |  |
| Rarely |  | 273 (19%) | 155 (20%) | 118 (17%) |  |
| **Vaccine** | 1,474 |  |  |  | 0.235 |
| Johnson & Johnson |  | 80 (5.4%) | 37 (4.9%) | 43 (6.0%) |  |
| Moderna |  | 427 (29%) | 219 (29%) | 208 (29%) |  |
| Other |  | 6 (0.4%) | 3 (0.4%) | 3 (0.4%) |  |
| Oxford-AstraZeneca |  | 105 (7.1%) | 65 (8.6%) | 40 (5.6%) |  |
| Pfizer-BioNTech |  | 856 (58%) | 436 (57%) | 420 (59%) |  |
| ^1^n (%); Median (Q1, Q3) for continuous variables | | | | | |
| ^2^Wilcoxon rank-sum test for continuous variables; Chi-square test for categorical variables | | | | | |
