## Supplementary Information 5 for "Menstrual cycle phase and its association with COVID-19 vaccine side effects and subsequent infection: A study of period tracking app users"

***S.I.5. Association of cycle phase at vaccination with any reported side effects, sensitivity and sub group analysis results***

| **Sensitivity Analysis** | **OR (95% CI)** | **P-value** |
| --- | --- | --- |
| **Not Obese** | **1.48 (1.12-1.95)** | **0.006** |
| **Post-secondary education** | **1.37 (1.07-1.74)** | **0.01** |
| **No medical conditions** | **1.41 (1.07-1.86)** | **0.02** |
| **Never smokers** | **1.54 (1.16-2.03)** | **0.003** |
| **White only** | **1.32 (1.00-1.73)** | **0.05** |
| mRNA only | 1.25 (0.98-1.60) | 0.07 |
| Pfizer only | 1.34 (1.00-1.80) | 0.05 |
| **Complete Cases** | **1.34 (1.05-1.70)** | **0.02** |
| **No Copper Coil** | **1.40 (1.10-1.79)** | **0.007** |
| **Not menstruating** | **1.34 (1.04-1.73)** | **0.02** |
| **Not periovulatory** | **1.29 (1.00-1.65)** | **0.05** |
| **Not perimenstrual** | **1.45 (1.11-1.89)** | **0.006** |
| **Systemic side effects** | **1.33 (1.04-1.68)** | **0.02** |

Note: Significant results (p < 0.05) are shown in bold.
