## Supplementary Information 6 for "Menstrual cycle phase and its association with COVID-19 vaccine side effects and subsequent infection: A study of period tracking app users"

***S.I.6. Interaction test results: Cycle phase at vaccination and any reported side effects***

| **Interaction Variable** | **P-value** | **Significant (p<0.05)** |
| --- | --- | --- |
| **Age Group (per 10 years)** | **0.04** | **Yes** |
| Residence | 0.12 | No |
| Vax1 Type | 0.15 | No |
| Smoking Status | 0.25 | No |
| BMI Category | 0.36 | No |
| Income Category | 0.43 | No |
| Stress Level (IQR) | 0.62 | No |

Note: Significant interactions (p < 0.05) are shown in bold. P-values from likelihood ratio tests comparing logistic regression models with and without interaction terms.

Detailed Results for Significant Interactions

| **Interaction Variable** | **Interaction P-value** | **Term** | **OR (95% CI)** | **Term P-value** |
| --- | --- | --- | --- | --- |
| **Age Group (per 10 years)** | **0.04** | **Cycle Phase: Follicular** | **2.22 (1.40-3.51)** | **<0.001** |
|  |  | **Cycle Phase: Follicular × Age: 25-34 years** | **0.51 (0.28-0.91)** | **0.02** |
|  |  | **Cycle Phase: Follicular × Age: 35-44 years** | **0.51 (0.27-0.94)** | **0.03** |

Note: Only interactions with p < 0.05 are shown. Significant individual terms (p < 0.05) are shown in bold. OR = Odds Ratio .
