## Supplementary Information 7 for "Menstrual cycle phase and its association with COVID-19 vaccine side effects and subsequent infection: A study of period tracking app users"

***S.I.7. Association of cycle phase at vaccination with reported side effect severity, sub group and sensitivity analysis results***

| **Sensitivity Analysis** | **Severity Comparison** | **OR (95% CI)** | **P-value** |
| --- | --- | --- | --- |
| **Not Obese** | **Overall** | **1.27 (1.01-1.60)** | **0.04** |
| Post-secondary education | Overall | 1.22 (1.00-1.49) | 0.05 |
| No medical conditions | Overall | 1.25 (0.99-1.57) | 0.06 |
| **Never smokers** | **Overall** | **1.29 (1.03-1.62)** | **0.03** |
| White only | Overall | 1.19 (0.95-1.49) | 0.13 |
| mRNA only | Overall | 1.15 (0.93-1.41) | 0.20 |
| Pfizer only | Overall | 1.25 (0.97-1.62) | 0.09 |
| Complete Cases | Overall | 1.21 (0.99-1.47) | 0.06 |
| **No Copper Coil** | **Overall** | **1.24 (1.01-1.51)** | **0.04** |
| Not menstruating | Overall | 1.21 (0.98-1.49) | 0.07 |
| Not periovulatory | Overall | 1.17 (0.95-1.44) | 0.13 |
| **Not perimenstrual** | **Overall** | **1.29 (1.04-1.61)** | **0.02** |
| **Systemic side effects** | **Overall** | **1.22 (1.00-1.49)** | **0.05** |

Note: Significant results (p < 0.05) are shown in bold. OR = Odds Ratio. Ordinal logistic regression with cumulative logits comparing higher severity levels vs. all lower levels combined.
