## Supplementary Information 8 for "Menstrual cycle phase and its association with COVID-19 vaccine side effects and subsequent infection: A study of period tracking app users"

***S.I.8. Interaction test results: Cycle phase at vaccination and severity of reported side effects***

| **Interaction Variable** | **P-value** | **Significant (p<0.05)** |
| --- | --- | --- |
| Age Group (per 10 years) | 0.48 | No |
| BMI Category | 0.69 | No |
| Income Category | 0.85 | No |
| Residence | 0.90 | No |
| Smoking Status | 0.96 | No |
| Stress Level (IQR) | 0.98 | No |
| Vax1 Type | 0.98 | No |

Note: Significant interactions (p < 0.05) are shown in bold. P-values from likelihood ratio tests comparing ordinal logistic regression models with and without interaction terms.

Detailed Results for Significant Interactions

| Interaction Variable | Interaction P-value | Term | OR (95% CI) | Term P-value |
| --- | --- | --- | --- | --- |
| No significant interactions found | N/A | N/A | N/A | N/A |

Note: Only interactions with p < 0.05 are shown. Significant individual terms (p < 0.05) are shown in bold. OR = Odds Ratio from ordinal logistic regression .
