## Supplementary Information 9 for "Menstrual cycle phase and its association with COVID-19 vaccine side effects and subsequent infection: A study of period tracking app users"

***S.I.9. Association of cycle phase at vaccination with reported number of Side effects, subgroup analysis results***

| **Sensitivity Analysis** | **IRR (95% CI)** | **P-value** |
| --- | --- | --- |
| Not Obese | 1.11 (0.99-1.25) | 0.07 |
| Post-secondary education | 1.09 (0.99-1.21) | 0.07 |
| No medical conditions | 1.10 (0.98-1.23) | 0.11 |
| Never smokers | 1.11 (0.99-1.24) | 0.07 |
| White only | 1.07 (0.96-1.20) | 0.20 |
| mRNA only | 1.04 (0.94-1.16) | 0.42 |
| Pfizer only | 1.11 (0.97-1.27) | 0.13 |
| Complete Cases | 1.08 (0.98-1.19) | 0.14 |
| No Copper Coil | 1.10 (0.99-1.21) | 0.08 |
| Not menstruating | 1.08 (0.97-1.20) | 0.16 |
| Not periovulatory | 1.07 (0.97-1.19) | 0.19 |
| Not perimenstrual | 1.10 (0.98-1.23) | 0.09 |
| Systemic side effects | 1.10 (0.99-1.22) | 0.07 |

Note: Significant results (p < 0.05) are shown in bold. IRR = Incidence Rate Ratio.
