## Supplementary Information 11 for "Menstrual cycle phase and its association with COVID-19 vaccine side effects and subsequent infection: A study of period tracking app users"

S.I. 11: COVID-19 infection risk by menstrual cycle phase at first vaccination: Cox proportional hazards regression results

| **Characteristic**^1^ | **HR**^1^ | **95% CI**^1^ | **p-value**^1^ |
| --- | --- | --- | --- |
| Cycle Phase (14-day luteal) |  |  |  |
| Luteal | — | — |  |
| Follicular | 0.73 | 0.47, 1.13 | 0.2 |
| Second Vaccine Status | 0.39 | 0.22, 0.70 | **0.001** |
| Age Group (per 10 years) |  |  |  |
| 25-34 | — | — |  |
| 18-24 | 1.22 | 0.72, 2.04 | 0.5 |
| 35-44 | 0.53 | 0.30, 0.93 | **0.026** |
| Medical Condition(s): Yes (vs No) |  |  |  |
| No medical condition | — | — |  |
| Has medical condition(s) | 1.13 | 0.69, 1.86 | 0.6 |
| ^1^HR = Hazard Ration. Bold valued indicate p <0.05. Cox proportional hazards model. N = 1474Events N =82; AIC = 1121.5; Wald test = 20.228; Wald test P-value = 0.001; Log-Likelihood = -555.772; LR test =19.063; C-index =0.641; R-squared =0.331 | | | |
| Abbreviations: CI = Confidence Interval, HR = Hazard Ratio | | | |
