## Supplementary Information 12 for "Menstrual cycle phase and its association with COVID-19 vaccine side effects and subsequent infection: A study of period tracking app users"

***S.I.12. Cox Model Stepwise Selection Summary***

| **Model** | **Variables Added** | **Events/Param** | **AIC** | **C-Index** | **Influential Obs** | **Max Change %** | **Stability** |
| --- | --- | --- | --- | --- | --- | --- | --- |
| Minimal | Base model | 27.3 | 1,126.3 | 0.615 | 41 | 100.0 | Unstable |
| Plus_PEC | Pre-existing conditions | 20.5 | 1,128.1 | 0.619 | 56 | 100.0 | Unstable |
| Plus_BMI | BMI category | 13.7 | 1,131.5 | 0.623 | 62 | 100.0 | Unstable |
| Plus_Vax | Vaccination status | 16.4 | 1,121.5 | 0.641 | 59 | 192.1 | Unstable |
