## Supplementary Information 13 for "Menstrual cycle phase and its association with COVID-19 vaccine side effects and subsequent infection: A study of period tracking app users"

***S.I. 14. 18-24 Young Age Group (18-24) vs Full Sample vaccine outcome comparisons within each menstrual cycle phase***

|  | **Luteal Phase Comparison** | | | **Follicular Phase Comparison** | | |
| --- | --- | --- | --- | --- | --- | --- |
| **Characteristic** | **Full Sample**  N = 714^1^ | **Young (18-24)**  N = 169^1^ | **p-value**^2^ | **Full Sample**  N = 760^1^ | **Young (18-24)**  N = 175^1^ | **p-value**^2^ |
| Presence of Side effects | 502 (70%) | 97 (57%) | 0.001 | 577 (76%) | 131 (75%) | 0.8 |
| Severity of Side effects |  |  | 0.012 |  |  | 0.9 |
| None | 212 (30%) | 72 (43%) |  | 183 (24%) | 44 (25%) |  |
| Weak | 324 (45%) | 59 (35%) |  | 386 (51%) | 92 (53%) |  |
| Moderate | 123 (17%) | 27 (16%) |  | 111 (15%) | 23 (13%) |  |
| Severe | 55 (7.7%) | 11 (6.5%) |  | 80 (11%) | 16 (9.1%) |  |
| COVID-19 infection | 46 (6.4%) | 12 (7.1%) | 0.8 | 36 (4.7%) | 11 (6.3%) | 0.4 |
| Time to COVID-19 (days) | 164 (107, 207) | 197 (153, 227) | 0.2 | 200 (140, 237) | 186 (71, 235) | 0.5 |
| ^1^P-values compare Young Age Group (18-24) vs Full Sample within each cycle phase | | | | | | |
| ^2^Pearson's Chi-squared test; Wilcoxon rank sum test | | | | | | |
