## Supplementary Information 14 for "Menstrual cycle phase and its association with COVID-19 vaccine side effects and subsequent infection: A study of period tracking app users"

***S.I. 13. Alternative cycle phase estimation model results***

| **Cycle Phase** | **Analysis** | **Effect** | **Effect (95% CI)** | **P-value** | **AIC** |
| --- | --- | --- | --- | --- | --- |
| **Standard (14-day luteal)** | **Presence** | **OR** | **1.35 (1.07-1.70)** | **0.013** | **1707.7** |
| Standard (14-day luteal) | Severity | OR | 1.21 (1.00-1.46) | 0.056 | 3587.1 |
| Standard (14-day luteal) | Count | RR | 1.08 (0.98-1.19) | 0.109 | 5779.1 |
| Standard (14-day luteal) | Time to COVID | HR | 0.73 (0.17-3.05) | 0.158 | 1121.5 |
| Standard (12-day luteal) | Presence | OR | 1.22 (0.96-1.54) | 0.099 | 1711.3 |
| Standard (12-day luteal) | Severity | OR | 1.15 (0.95-1.40) | 0.156 | 3588.8 |
| Standard (12-day luteal) | Count | RR | 1.05 (0.95-1.16) | 0.345 | 5780.8 |
| Standard (12-day luteal) | Time to COVID | HR | 0.74 (0.17-3.19) | 0.180 | 1121.8 |
| **Standard (13-day luteal)** | **Presence** | **OR** | **1.29 (1.02-1.63)** | **0.032** | **1709.4** |
| Standard (13-day luteal) | Severity | OR | 1.21 (1.00-1.47) | 0.053 | 3587.0 |
| Standard (13-day luteal) | Count | RR | 1.07 (0.97-1.18) | 0.151 | 5779.6 |
| Standard (13-day luteal) | Time to COVID | HR | 0.73 (0.17-3.02) | 0.149 | 1121.5 |
| Age-adjusted | Presence | OR | 1.21 (0.96-1.53) | 0.113 | 1711.5 |
| Age-adjusted | Severity | OR | 1.15 (0.94-1.39) | 0.168 | 3588.9 |
| Age-adjusted | Count | RR | 1.05 (0.96-1.16) | 0.299 | 5780.6 |
| Age-adjusted | Time to COVID | HR | 0.76 (0.17-3.33) | 0.206 | 1122.0 |
| Cycle-length-adjusted | Presence | OR | 1.22 (0.97-1.55) | 0.089 | 1711.1 |
| Cycle-length-adjusted | Severity | OR | 1.16 (0.96-1.41) | 0.133 | 3588.5 |
| Cycle-length-adjusted | Count | RR | 1.05 (0.95-1.16) | 0.308 | 5780.6 |
| Cycle-length-adjusted | Time to COVID | HR | 0.74 (0.17-3.16) | 0.175 | 1121.7 |

Note: OR = Odds Ratio, RR = Rate Ratio, HR = Hazard Ratio. Significant results (p < 0.05) in bold.
